# Yellow fever re-emergence in Tolima, Colombia 2024-25: an eco-epidemiological study

**DOI:** 10.64898/2026.01.16.26344301

**Authors:** Seth D. Judson, Nicole Stephan, Sofia Machuca, Andrés F. Henao-Martínez, Gabriel Parra-Henao, Ricardo Vivas, Jose Fair Alarcón-Robayo, Mauricio Javier Vera Soto, Hernán Vargas

**Affiliations:** Division of Infectious Diseases, Department of Medicine, David Geffen School of Medicine at University of California, Los Angeles, USA; Division of Infectious Diseases, Department of Medicine, University of Colorado Anschutz Medical Campus, Aurora, USA; Centro de Investigación en Salud para el Trópico, Universidad Cooperativa de Colombia, Santa Marta, Colombia; Dirección de Salud Pública, Secretaria de Salud del Tolima, Colombia; Subdirección de Enfermedades Transmisibles, Ministerio de Salud y Protección Social, Bogotá, Colombia; Grupo de investigación clínica y medicina traslacional, Hospital Federico Lleras Acosta, Ibagué, Colombia; Grupo de inmunologia molecular, Universidad del Quindío, Armenia, Colombia

**Author notes:** Corresponding author: Seth D. Judson, Division of Infectious Diseases, 924 Westwood Blvd #200, Los Angeles, CA, USA.

## Abstract

**Background:** Yellow fever virus (YFV) is transmitted by mosquitoes among humans and non-human primates (NHPs) in South America. The 2024-25 yellow fever (YF) outbreak was notable for its spread into new areas, including the department of Tolima in Colombia’s Andean region. We investigated the eco-epidemiology of human and NHP YF cases to understand the patterns and factors associated with the Tolima outbreak.

**Method:** We collected spatiotemporal, sociodemographic, and mortality data on human YF cases in Tolima, as well as the locations of deceased NHPs with YF. We then conducted exploratory, descriptive, and spatial statistical analyses to identify risk factors and hotspots for YF. We then used inferential spatiotemporal modeling to compare ecological and sociodemographic factors.

**Results:** From September 2024 to October 2025, 116 human and 53 non-human primate YF cases were detected in Tolima. Among the human cases, 77% were male (89/116), and the median age was 47 (interquartile range 34-63). Of these cases, 45 (39%) died, and older age was associated with increased odds of death (adjusted odds ratio, 1.43 per 10 years; 95% confidence interval, 1.18-1.78). There was significant clustering among human and NHP cases. Higher rainfall and poverty/deprivation were associated with increased YF incidence at the neighborhood level, and rainfall during the outbreak was above average due to La Niña.

**Conclusions:** These findings demonstrate significant mortality from YF, which re-emerged in Tolima after nearly a century. Rural populations with greater ecological risk and poverty/deprivation could benefit from targeted vaccination strategies for YF.

**Summary:** Yellow fever re-emerged in Tolima, Colombia after nearly a century, causing mortality among humans and non-human primates. This outbreak occurred primarily in rural areas with low vaccination coverage, highlighting the need for preparedness in areas with similar risk factors.

## Introduction

During 2024-2025, a yellow fever (YF) outbreak in South America expanded into new regions, causing global concern. Yellow fever is a viral hemorrhagic fever caused by a mosquito-borne flavivirus, yellow fever virus (YFV). In South America, the Amazon region is often considered the primary endemic area for YFV circulation. However, in October 2024, YF cases were reported in Tolima, Colombia, a department in the Andean region, which had no known YF cases in the 21st century and low YF vaccination coverage [1–7]. Concerning features of the 2024-2025 Tolima outbreak included substantial morbidity and mortality, as well as potential peri-urban cases [4,7].

The majority of both historical and recent YF cases in South America have been reported in Brazil, Bolivia, Colombia, Peru, and Ecuador [8,9]. A significant YF outbreak occurred in Brazil in 2016-2018, notable for the re-emergence of YF in the Atlantic forest region and proximity to major cities. In Colombia, there have been no major YF outbreaks since 2005, aside from sporadic cases [10]. Thus, the YF outbreak in Tolima is highly concerning and part of a recent trend of YF outbreaks occurring among different ecoregions during the past decade.

Yellow fever virus circulates via two transmission cycles in the Americas—the sylvatic and urban cycles [8]. The sylvatic cycle involves non-human primate (NHP)-to-mosquito-to-human transmission near forested areas via primarily *Haemagogus* and *Sabethes* mosquitoes. Some new-world NHPs, especially howler monkeys (*Alouatta spp.*), are susceptible to severe disease and mortality from the sylvatic cycle of YFV [11]. Yellow fever outbreaks in South America have almost exclusively been attributed to the sylvatic cycle since the 1940s, in the context of urban mosquito control and YF vaccination efforts [7].

Given that the severe form of YF has an estimated case fatality ratio (CFR) of 39% [12], and there are no licensed therapeutics for YF, prevention is key. Understanding the eco-epidemiology of the YF outbreak in Tolima could yield novel insights into risk factors and spatiotemporal patterns of YF, enabling targeted preventive measures such as vaccination and vector control. While multiple hypotheses have been generated surrounding the drivers of the Tolima outbreak [4,5], spatial statistical and modeling analyses could help understand these factors. Therefore, the goals of our study were to (1) characterize the epidemiology and disease burden of YF in Tolima, (2) analyze spatiotemporal dynamics of YF spread, and (3) identify ecological and sociodemographic risk factors associated with YF incidence.

## Methods

### Setting

The administrative structure of Colombia comprises 32 departments, divided into municipalities, which are further divided into the lowest level, called veredas (neighborhoods). Tolima is a department in Colombia’s central-west region and has 47 municipalities. These municipalities include many rural areas and dispersed settlements, as well as urban areas such as the capital city, Ibagué.

Tolima has varied topography, including two major branches of the Andes Mountains separated by the Magdalena River Valley [13]. This altitudinal variation contributes to diverse ecosystems in Tolima, including lowland riparian forests, mid-altitude Andean forests, and high-Andean forest and páramos [13]. Agriculture is the most important economic sector and has been associated with deforestation as well as increased seasonal human mobility [2]. Additionally, there has been an increase in refugees and migration into the region from Venezuela, as well as armed conflict in neighboring departments of Colombia [2].

Historical YF outbreaks in Tolima occurred from 1830-1940 (Fig. S1) [14–16]. Given the absence of further cases in the region, Tolima was considered a low-risk zone for YF and vaccination was optional. In 2005, routine YF vaccination in Tolima was implemented only for infants. An estimated 54% of adults had been vaccinated prior to the outbreak [2].

### Human and non-human primate cases

For the 2024-2025 YF outbreak, we analyzed confirmed human YF cases in Tolima. We included cases from the earliest confirmed case (based on symptom onset), which occurred September 8^th,^ 2024, to October 31^st,^ 2025. Cases were confirmed at the Laboratorio Nacional de Referencia, del Instituto Nacional de Salud de Colombia via reverse transcription polymerase chain reaction (RT-PCR) [17]. Demographic data, date of symptom onset, mortality status, and location of residence were collected on all cases. Additionally, data were collected by the Corporación Autonoma Regional del Tolima on deceased NHPs in Tolima that were confirmed to have YFV infection post-mortem via RT-PCR and/or histopathology. The testing of NHPs began in February 2025, and the locations of these NHPs were collected, including municipalities and veredas, and georeferenced GPS coordinates, when available.

### Statistical analysis

Descriptive statistics of confirmed human YF cases were calculated in R version 4.5.2. We conducted univariable and multivariable logistic regression to compare the associations between age and sex with mortality status. To analyze areal-level spatial relationships, we aggregated cases to the municipality and vereda levels and assessed spatial autocorrelation using Moran’s I (Figs. S2, S3). For point-pattern analysis, the geographic coordinates of residences of confirmed human cases and the locations of infected deceased NHPs were obtained, and the K and L functions were calculated in R (Figs. S4-S7).

### Ecological and sociodemographic factors

Using a conceptual framework, we identified key covariates potentially associated with YF incidence in Tolima (Fig. S8). These covariates were selected based on whether they had previously been associated with YF in other regions, as well as on discussions with public health officials in Tolima [18–20]. The sources and spatiotemporal resolution for these covariates are shown in Table S1. We included ecological covariates (precipitation, temperature, elevation, vegetation, and land cover) and sociodemographic covariates (population density, poverty/deprivation, and built-up area) (Tables S1-S3). We also assessed the species distribution of NHPs in Tolima using the IUCN Redlist (Fig. S9) [21]. We conducted an initial exploratory statistical analysis by comparing each covariate between veredas that reported YF cases and those that did not (Appendix, section 5). Because granular vaccination coverage data prior to the outbreak were unavailable, vaccination was not included in the analysis.

### Spatiotemporal modeling approach

We used a Bayesian inferential modeling approach to identify which covariates were associated with YF incidence in Tolima from 2024 to 2025. First, a correlation matrix was used to exclude covariates that had a correlation coefficient of >|0.7| (Appendix, section 2). We then developed four types of Bayesian hierarchical spatiotemporal models using integrated nested Laplace approximation using the R-INLA package. Model I includes fixed effects only. Model II includes fixed effects and spatially structured and unstructured random effects using a Besag-York-Mollié (BYM2) model. Model III adds an unstructured space-time interaction term to capture unexplained temporal variation at the vereda level. Model IV is a full spatiotemporal model that additionally includes a temporally structured random effect, modeled as a first-order random walk. We compared the best fitting of these models to a baseline model with only random effects. Model fit was assessed using the deviance information criterion (DIC) and the Watanabe-Akaike information criterion (WAIC). All covariates were standardized, and we assessed lag values of monthly temperature, precipitation, and NDVI ranging from 1 to 3 months to identify the optimal lag to include in the models. To account for overdispersion, monthly YF cases were modeled as a negative binomial process:

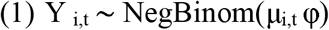

where φ is the overdispersion parameter and Y _i,t_ is the observed number of YF cases in vereda *i* during month *t*, while μ_i,t_ is the expected mean number of cases in *i* at *t,* modeled in the spatiotemporal model as the log-incidence rate:

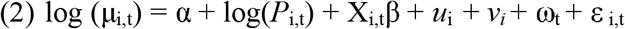

where α is the intercept, log(*P*_i,t_) is an offset to adjust case counts by the logarithm of the population, and X is a matrix of covariates with regression coefficients β. The spatial component follows the Besag-York-Mollié (BYM2) model, where *u_i_* is the spatially structured random effect (conditional autoregressive), and *v_i_* is the unstructured (i.i.d.) vereda-level random effect. The temporal trend, ω_t,_ is modeled as a first-order random walk, and ε _i,t_ is an unstructured space-time interaction.

### Patient consent statement

This study was deemed exempt from institutional review board review (IRB) by the University of California, Los Angeles IRB, and informed consent was waived, as it involved only secondary analyses of data previously collected for public health purposes.

## Results

### Human yellow fever cases

From 2024 to 2025, 116 laboratory-confirmed cases of YF were identified in Tolima (Table 1). Among these cases, 45 (39%) died, all of whom were unvaccinated for YF. Of the nine YF cases that were previously vaccinated, per case investigations these were not attributed to vaccine-associated viscerotropic disease. Overall these cases in Tolima represented 85% (116/136) of all YF cases reported in Colombia from January 1^st^, 2024, to October 31^st^, 2025 [9].

**Table 1.**
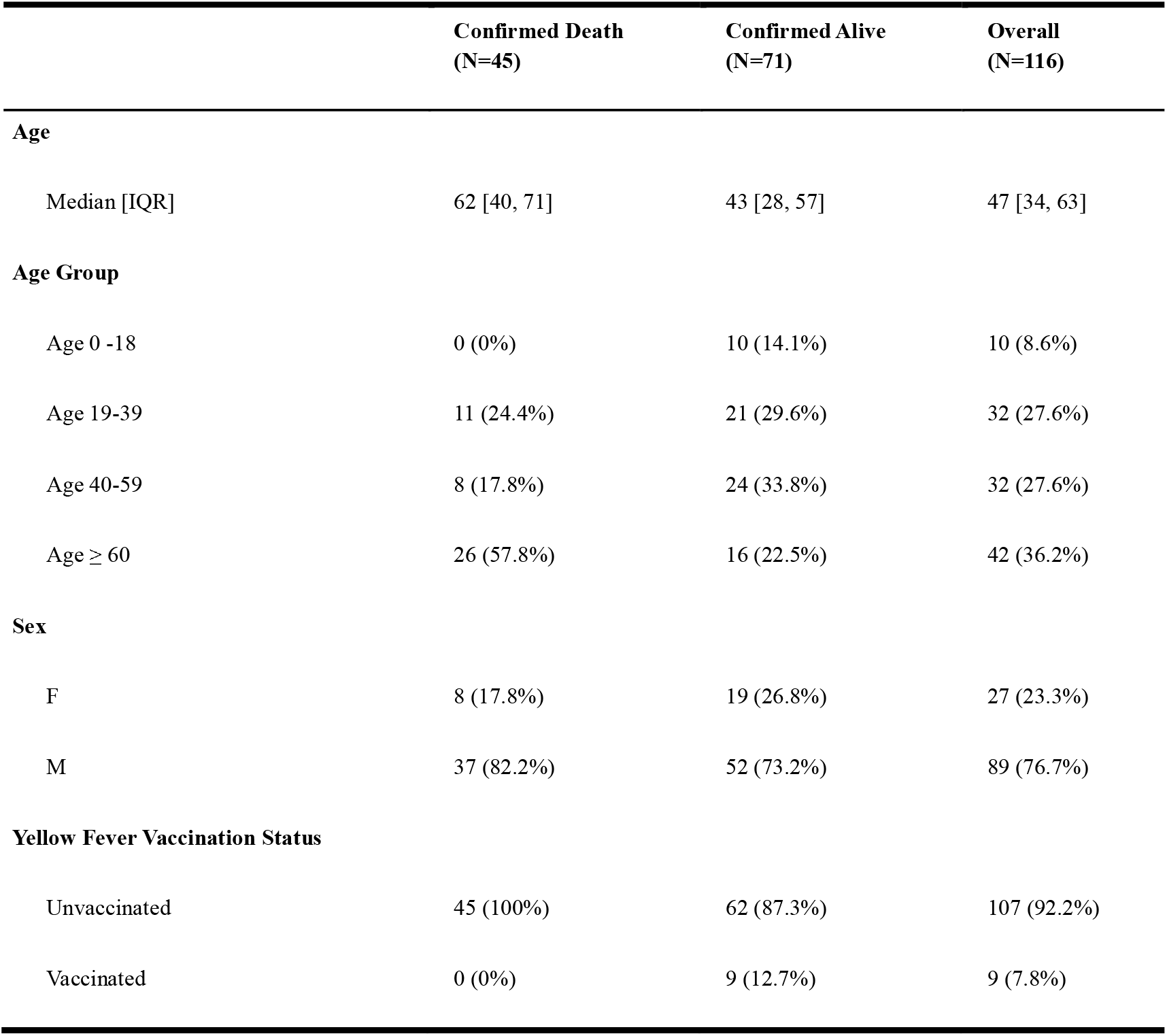
Yellow Fever Cases in Tolima, Colombia, September 2024 – October 2025.

|  | Confirmed Death<br>(N=45) | Confirmed Alive<br>(N=71) | Overall<br>(N=116) |
| --- | --- | --- | --- |
| Age |  |  |  |
| Median [IQR] | 62 [40, 71] | 43 [28, 57] | 47 [34, 63] |
| Age Group |  |  |  |
| Age 0 -18 | 0 (0%) | 10 (14.1%) | 10 (8.6%) |
| Age 19-39 | 11 (24.4%) | 21 (29.6%) | 32 (27.6%) |
| Age 40-59 | 8 (17.8%) | 24 (33.8%) | 32 (27.6%) |
| Age ≥ 60 | 26 (57.8%) | 16 (22.5%) | 42 (36.2%) |
| Sex |  |  |  |
| F | 8 (17.8%) | 19 (26.8%) | 27 (23.3%) |
| M | 37 (82.2%) | 52 (73.2%) | 89 (76.7%) |
| Yellow Fever Vaccination Status |  |  |  |
| Unvaccinated | 45 (100%) | 62 (87.3%) | 107 (92.2%) |
| Vaccinated | 0 (0%) | 9 (12.7%) | 9 (7.8%) |

The majority of YF cases were male (77%, 89/116), and the overall median age of cases was 47 [interquartile range (IQR) 34-63]. A higher proportion of deaths occurred among those ≥ 60 years old (58%, 26/45). In univariable and multivariable logistic regression analyses, adjusting for sex, age was associated with increased odds of death (adjusted odds ratio, aOR, 1.43 per 10 years; 95% confidence interval, 1.18-1.78) (Table S5).

The initial YF cases were detected in the southeastern municipalities, with the highest incidence in Cunday, followed by Villarrica, Prado, Purificacion, and Dolores. Subsequently, cases were detected in the central part of the department with single cases in Ibagué, El Espinal, and Valle de San Juan, as well as a single case in the northern municipality of Palocabildo. Multiple YF cases were then detected in the southern municipalities of Ataco, Chaparral, and Rioblanco. There were 74 veredas with YF cases. The incidence of YF cases by municipality and vereda are shown in Figure 1.

**Figure 1.**
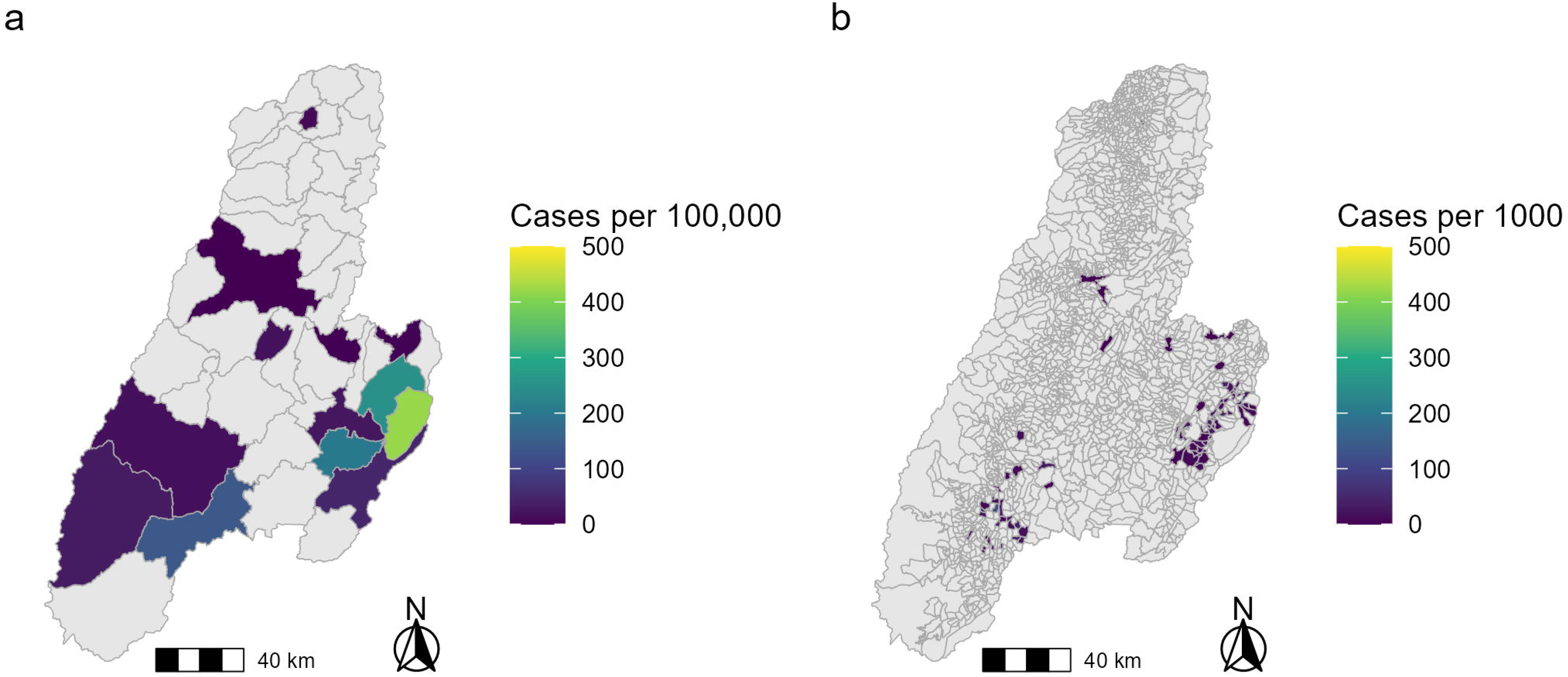
Incidence Rate of Human Yellow Fever Cases in Tolima, Colombia 2024-2025. The incidence rate of human yellow fever cases from September 8^th^ 2024 to October 31^st^, 2025 in Tolima are shown aggregated to the (a) municipality level and (b) vereda level. Alt text: Two maps of the department of Tolima, Colombia showing the incidence of yellow fever cases by municipality and vereda

### Non-human primate yellow fever cases

Of 53 confirmed NHP cases with vereda locations, 49 individuals had georeferenced coordinates available (Fig. S4). The NHP cases occurred in the following municipalities (in order of highest incidence): Chaparral, Ataco, Planadas, Cunday, Purificacion, Villarrica, Rioblanco, San Antonio, and Prado. The genera of NHPs infected included *Alouatta* and *Aotus*.

### Spatiotemporal analysis

Human YF cases occurred between September 8^th^, 2024, and October 8^th^, 2025. Non-human primate cases were detected from February 10^th^, 2025 to October 1^st^, 2025. Figure 2 depicts the epidemic curve with the timing of human and NHP cases.

**Figure 2.**
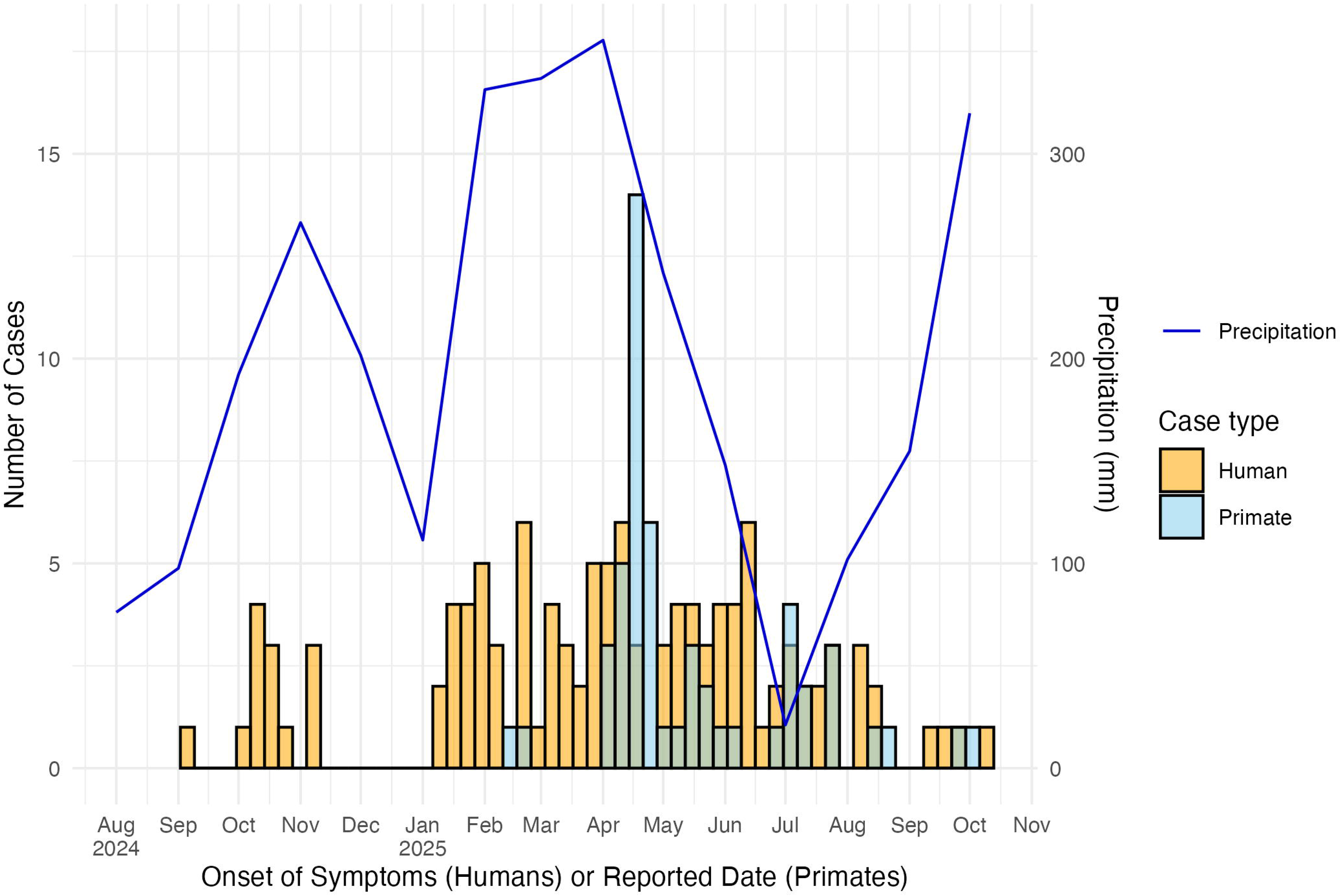
Epidemic Curve of Human and Primate Yellow Fever Cases in Tolima, 2024-2025. The histogram depicts yellow fever cases by week of symptom onset for humans (orange) and reported date of death for non-human primates (blue). The line graph shows the overall mean monthly precipitation in Tolima, given the significant association between YF incidence in Tolima and precipitation. Alt text: A histogram of weekly yellow fever cases in humans and non-human primates with a blue line for precipitation

Significant spatial autocorrelation among human YF incidence rates was found via Global Moran’s I at the municipality level (*I* = 0.24, p < 0.001), and weaker spatial autocorrelation at the vereda level (*I* = 0.03, p <0.001). Neighboring municipalities and veredas with high YF incidence rates (hotspots) were identified using Local Moran’s I (Figs. S2, S3). These hotspots occurred in two areas in southern and southeastern Tolima.

The spatial patterns of human vs NHP YF cases are shown in Figure 3. Human YF cases were found to significantly cluster based on the L function, with clustering strongest at 23.9 km (Fig. S6). Non-human primate YF cases were found to form statistically significant clusters with human cases, with clustering strongest at 25 km (Fig. S7).

**Figure 3.**
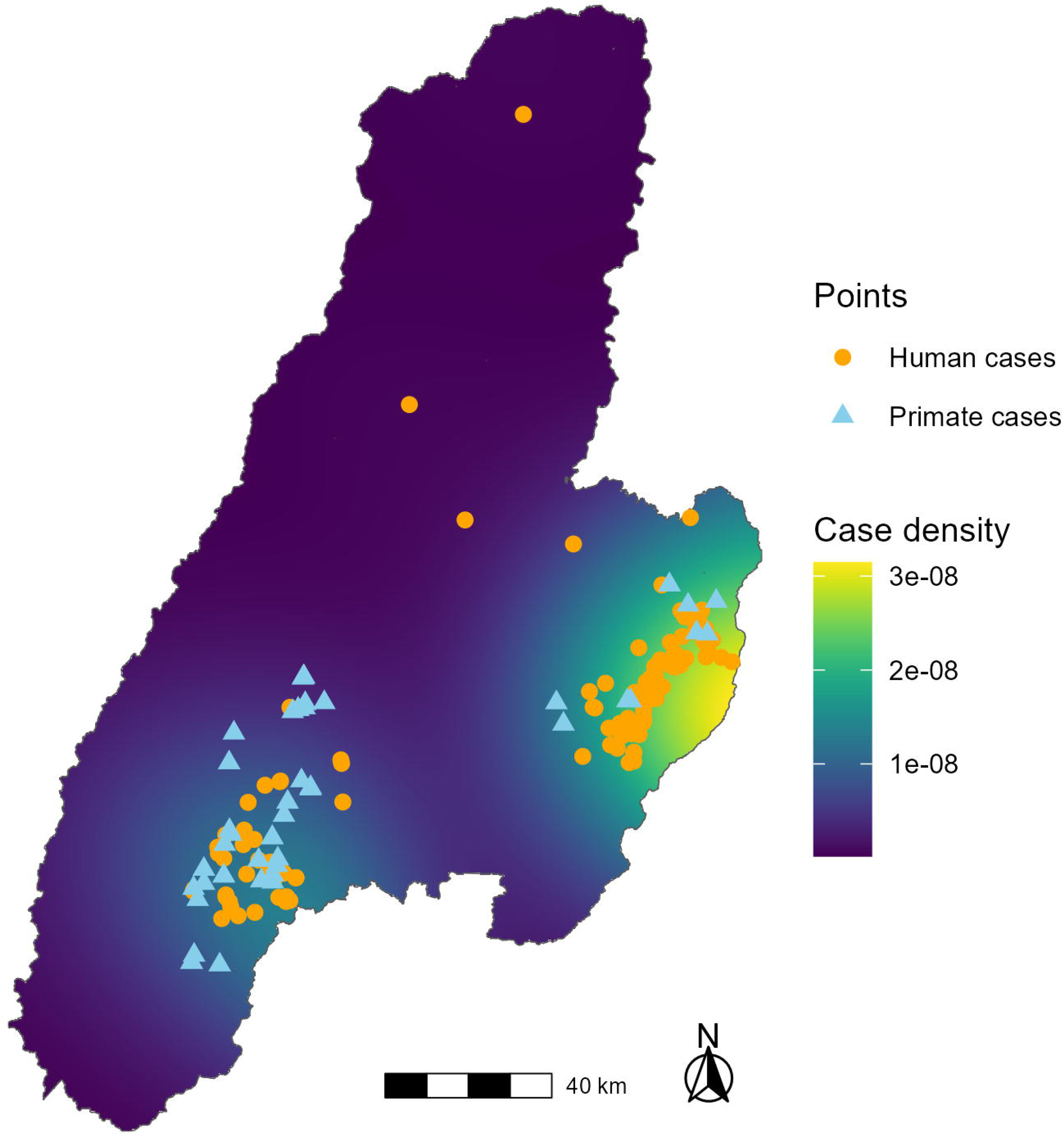
Kernel Density of Human Yellow Fever Cases in Tolima, Colombia 2024-2025. The georeferenced locations of human and non-human primate yellow fever cases in Tolima are shown. The L function was used to calculate the distance at which maximal human case clustering occurred (23.9 km). Thus, 23.9 km was selected for the bandwidth of the kernel density estimate to depict the intensity of clustering. Alt text: A map with points for locations of human and non-human primate cases with clustering intensity highlighted

### Exploratory analysis of ecological and sociodemographic factors

Figure 4 shows the distribution of ecological and sociodemographic covariates in Tolima. Monthly mean precipitation (Fig. S10-12) and temperature (Fig. S13-15) were higher in veredas with YF cases compared to those without cases (p-value <0.001). Yellow fever cases occurred at a mean elevation of 1066 meters (range 318-2190 meters) (Table S3), and mean elevation was lower in veredas with cases (p = 0.007, Fig S16). The highest proportion of YF cases occurred in a mosaic of agriculture and pasture habitat (71%, 82/116), followed by forest (23%, 27/116), infrastructure (3%, 4/116), other non-vegetated area (1%, 1/116), and other non-forest formation (2%, 2/116) (Table S4, Fig. S17). Four YF cases occurred in cities/dense towns (including Ibagué) based on urban built-up surface classification. There were no statistically significant differences in overall monthly mean NDVI, population density, poverty/deprivation index, or built-up surface between veredas with YF cases and those without cases (Figs. S18-S21).

**Figure 4.**
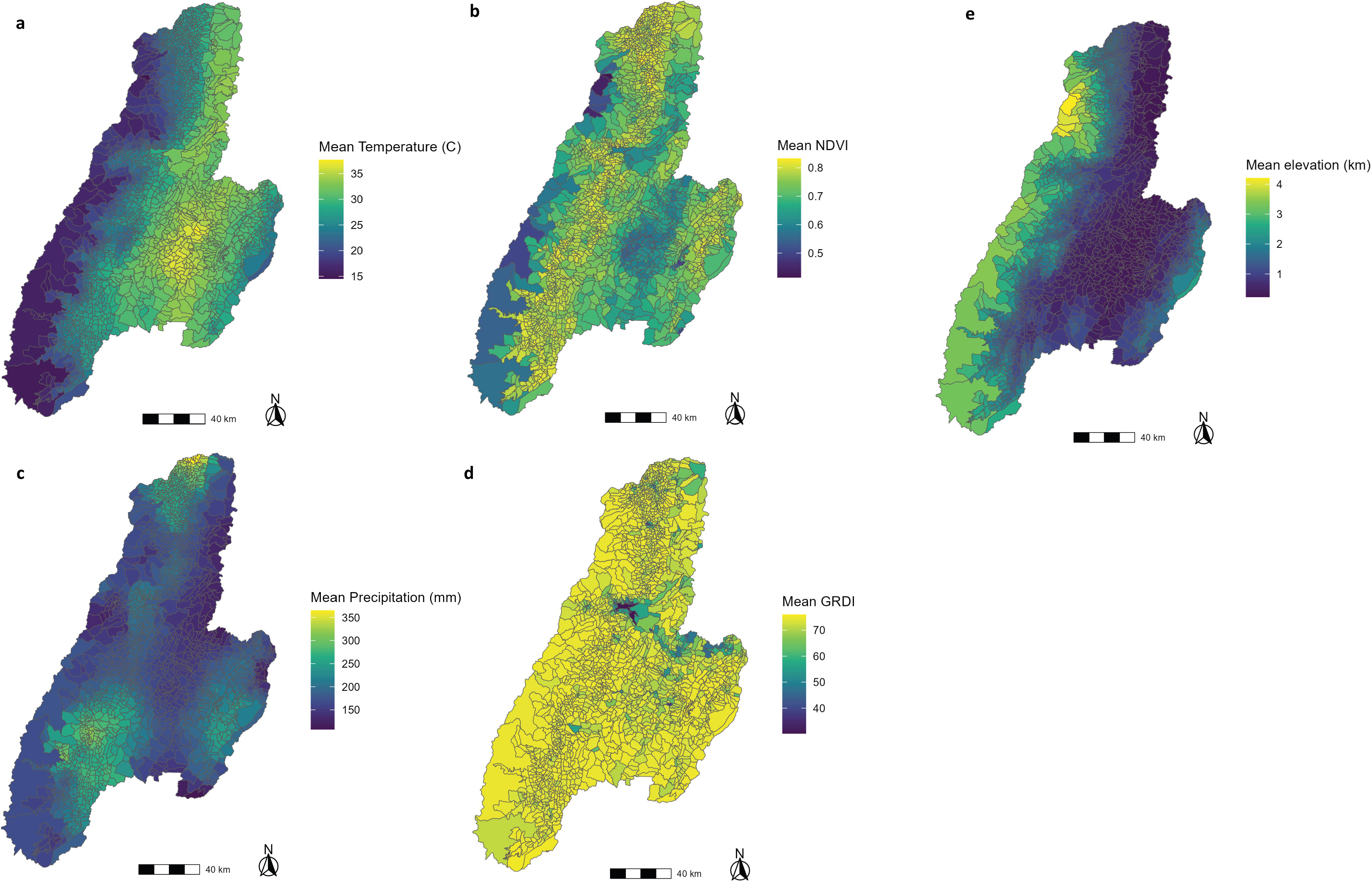
Ecological and Sociodemographic Covariates in Tolima, Colombia 2024-2025. The multipanel figure depicts the vereda-level mean (a) temperature 2024-2025, (b) Normalized Difference Vegetation Index (NDVI) 2024-2025, (c) precipitation 2024-2025, (d) Global Gridded Relative Deprivation Index 2020, and (e) elevation. Alt text: Maps of the mean temperature, vegetation, precipitation, deprivation/poverty, and elevation in Tolima, Colombia.

### Spatiotemporal modeling results

Comparing our Bayesian hierarchical spatiotemporal modeling results, the spatiotemporal model with standardized covariates and a 3-month lag for precipitation, temperature, and NDVI had optimal performance, as indicated by the lowest WAIC and DIC (Table S5). For this model, the incidence rate ratio (IRR) for precipitation was 2.32 (95% credible interval 1.62 – 3.47), temperature 1.25 (95% credible interval 0.79 – 1.98), poverty/deprivation 1.74 (95% credible interval 1.34 – 2.25), and NDVI 1.37 (95% credible interval 0.99 – 1.89). All models identified an increased incidence rate of YF associated with higher mean monthly precipitation and higher mean poverty/deprivation at the vereda level (Table S6). The full model had better fit and spatial precision than the baseline model, and structured spatial effects contributed most to the variance (Table S7).

## Discussion

The 2024-2025 YF outbreak in Tolima was the largest regional YF outbreak during this period in South America. While YF cases had occurred in Tolima from 1830 to 1940, there were no reported cases in this department for nearly a century. Overall, the epidemiological features of this outbreak are reminiscent of other YF outbreaks in South America, but with key differences. As with other YF outbreaks in South America, the highest proportion of YF cases occurred among middle-aged males, a demographic that includes agricultural workers most likely to be exposed to the sylvatic cycle. However, there was also a high proportion of patients aged 60 years or older with YF (36%), who had a very high CFR (58%), and increased age was found to be significantly associated with mortality. The elderly population was particularly vulnerable during this outbreak because Tolima was considered a low-risk department for YF and had low vaccination coverage.

Additionally, the overall CFR was very high (39%), which is consistent with global estimates for severe YF, indicating that most likely severe YF cases were detected in Tolima, whereas mild or asymptomatic cases were underascertained. It is estimated that an average of 12% of YF cases are severe [22], suggesting that roughly 1000 YF cases could have occurred in Tolima. The incidence of YF was higher among veredas with greater poverty/deprivation in Tolima, particularly in rural areas. These communities likely have less access to healthcare and lower levels of education. Residents of rural areas are also more likely to be exposed to sylvatic YF vectors due to work activities and proximity to forest borders. Since Tolima was classified as a low-risk department for YF, vaccination remained optional for adults, leading to a significant susceptible population. These findings emphasize the critical importance of immunization for YF (both routine childhood vaccination and mass vaccination campaigns) in areas where YF may circulate, including the Andean region.

We identified clustering of human and NHP cases, particularly around two subsequent foci in the southeastern and southern regions of Tolima. This clustering was strongest between 20 and 30 km for humans and NHPs, and human cases primarily occurred in transitional habitats. We found that precipitation was associated with YF incidence in the Tolima outbreak. La Niña that occurred in late 2024 through 2025 might have been a factor contributing to the above-average rainfall during the outbreak.

Further considering the ecology of the YF outbreak in Tolima, two primary NHP genera were diagnosed with YF post-mortem (*Alouatta* and *Aotus*). The majority of YF cases occurred among *Alouatta seniculus*, which are highly susceptible to YF and exhibit high mortality [11]. *Alouatta spp.* are widespread and are able to survive in fragmented landscapes, including Andean forests that have undergone deforestation [23]. *Aotus spp.* also often develop fatal infection and high viremia with YFV and can live in peri-urban forest fragments [11,24,25]. Therefore both *Alouatta* and *Aotus* were involved in the Tolima outbreak, but to what degree is less clear. Other species present in Tolima include *Sapajus apella*, *Cebus versicolor*, and *Lagothrix lagothricha,* which are thought to develop less severe infection from YFV. However, both *Cebus* and *Lagothrix* were recently found to develop fatal YFV infection under captive conditions in Colombia [26]. Multiple epizootic YF outbreaks among NHPs were reported during 2024-2025, including the department of Huila which borders Tolima [27]. Given that there was not a national surveillance system for YF in NHPs in Colombia, deceased NHPs were tested in response to the YF outbreak in humans, explaining the lag between cases. Additionally, NHP cases were reported by date of death, making it difficult to infer whether NHP cases preceded, coincided with, or followed human transmission.

The majority of YF cases occurred in rural areas near forested habitats. Initial cases occurred near Bosque de Galilea Natural Park among individuals involved in agricultural and logging activities in the forest. Deforestation could increase habitat fragmentation for NHPs and bring humans into closer contact with YFV-infected NHPs and vectors. Such forest fragmentation has been considered to be a driver of YF spread in other regions [28].

While most YF cases occurred in rural areas, four individual cases were reported near urban areas, including one in the capital city of Ibagué. Additional epidemiological and entomological analyses are needed to determine where these peri-urban cases most likely acquired infection. To truly determine the ecological transmission cycles and origins of this outbreak, entomological and viral genomic data are needed.

There are multiple limitations to our analysis. We used the locations of residence of human YF cases as a proxy for where infection occurred. The degree of clustering between human and NHP cases could be an overestimate because NHP deaths are more likely to be recognized in more populated areas. To compare covariates with YF incidence and protect privacy, we aggregated cases to the areal level. This can lead to the Modifiable Areal Unit Problem (MAUP), a bias introduced by aggregating continuous geographic variables into discrete areas. However, we minimized this by aggregating to the vereda level, the smallest administrative unit with a mean area of 12 km^2^ per vereda (Appendix, section 4). While all covariates were aggregated to the vereda level for our analysis, monthly precipitation and temperature were at a coarser spatial resolution than the other covariates. Local downscaling techniques that account for cloud cover would be needed to obtain these covariates at higher resolution. While we were able to compare multiple factors from our conceptual framework, we were unable to directly assess the role of YF vaccination coverage due to a lack of granular data. In the future, including vaccination coverage, as well as data on human mobility (transhumance) and entomologic surveillance could help improve our understanding of transmission dynamics.

In summary, the re-emergence of YF in Tolima revealed critical lessons regarding YF eco-epidemiology and prevention. While this region had not experienced YF for nearly a century, it had the appropriate ecological conditions for the spread of YF. These conditions and a rural, unvaccinated elderly population with high levels of poverty and deprivation, fueled a substantial YF outbreak. Identifying other regions with similar ecological and sociodemographic risk factors is essential for targeted YF vaccination campaigns to prevent future outbreaks.

## Data availability

The individual line-level human data from this study contain protected health information and are restricted from public sharing. These data may be made available upon request, subject to approval from the Secretaría de Salud del Tolima, Colombia, and from institutional review boards. Municipality and vereda-level aggregate human data and NHP data from this study and R scripts for the analysis are available at: https://doi.org/10.6084/m9.figshare.31061575 and https://github.com/Judson-Lab/YF_Tolima

## Acknowledgements

Funding for this study was provided by the University of California, Los Angeles.

## Author contributions

Conceptualization: SDJ, AFH, GP, RV, HV; Data curation: SDJ, NS, SM, RV, HV; Formal Analysis: SDJ, NS, SM; Funding acquisition: SDJ; Methodology: SDJ, NS; Writing – original draft: SDJ; Writing – review & editing: SDJ, NS, SM, AFH, GP, RV, JFA, MJS, HV

## References

1. Yellow Fever: How three monkey deaths sparked a critical health alert in Colombia - PAHO/WHO | Pan American Health Organization [Internet]. [cited 2025 Nov 23]. Available from: https://www.paho.org/en/stories/yellow-fever-how-three-monkey-deaths-sparked-critical-health-alert-colombia

2. ACAPS Thematic Report - Colombia: Yellow fever outbreak in Tolima (05 February 2025) - Colombia | ReliefWeb [Internet]. 2025 [cited 2025 Nov 23]. Available from: https://reliefweb.int/report/colombia/acaps-thematic-report-colombia-yellow-fever-outbreak-tolima-05-february-2025

3. Forero-Delgadillo AJ, Morales-Olivera JA, Celis-Guzmán JF, et al. Colombian consensus on the care of critically ill patients with suspected or confirmed severe yellow fever. Lancet Reg Health - Am. 2025; 48:101144.

4. Cuéllar-Sáenz JA, Rodríguez-Morales AJ, Faccini-Martínez ÁA. Early Release - Reemergence of Yellow Fever, Magdalena Valley, Colombia, 2024–2025 - Volume 31, Number 12—December 2025 - Emerging Infectious Diseases journal - CDC. 2025 [cited 2025 Dec 12]; . Available from: https://wwwnc.cdc.gov/eid/article/31/12/25-1209_article

5. Alvarez-Moreno CA, Rodriguez-Morales AJ. Challenges of the current yellow fever outbreak in Colombia. The Lancet. Elsevier; 2025; 405(10497):2273.

6. Taylor L. WHO warns of “explosive” yellow fever outbreaks as disease spreads in Americas. BMJ. British Medical Journal Publishing Group; 2025; 389:r1106.

7. Parra-Henao G, Usme-Ciro JA, Fernández-Niño JA, Henao-Martínez AF. Yellow fever’s distressing return: a wake-up call for public health in the Americas. Ther Adv Infect Dis. 2025; 12:20499361251359017.

8. Monath TP, Vasconcelos PFC. Yellow fever. J Clin Virol. Elsevier; 2015; 64:160–173.

9. PAHO/WHO - Yellow Fever in Americas Region [Internet]. Dashboard PAHOWHO - Yellow Fever AMRO. [cited 2025 Nov 21]. Available from: https://shiny.paho-phe.org/yellowfever/

10. Sanchez-Rojas IC, Solarte-Jimenez CL, Chamorro-Velazco EC, et al. Yellow fever in Putumayo, Colombia, 2024. New Microbes New Infect. 2025; 64:101572.

11. Risk assessment on yellow fever virus circulation in endemic countries [Internet]. [cited 2023 Nov 7]. Available from: https://www.who.int/publications-detail-redirect/WHO-HSE-PED-CED-2014-2

12. Servadio JL, Muñoz-Zanzi C, Convertino M. Estimating case fatality risk of severe Yellow Fever cases: systematic literature review and meta-analysis. BMC Infect Dis. 2021; 21(1):819.

13. Hermelin M. Geomorphological Landscapes and Landforms of Colombia. In: Hermelin M, editor. Landsc Landf Colomb [Internet]. Cham: Springer International Publishing; 2016 [cited 2025 Nov 21]. p. 1–21. Available from: 10.1007/978-3-319-11800-0_1

14. Smith HH, Bevier G, Bugher JC. The Distribution of Yellow Fever in Colombia in Recent Years. Am J Trop Med Hyg. The American Society of Tropical Medicine and Hygiene; 1943; s1-23(5):505–522.

15. Quevedo V. Emilio. De los litorales a las selvas : la construcción del concepto de fiebre amarilla selvática, 1881-1938 /. 1. ed. Bogotá, Colombia : Universidad del Rosario,;

16. Gast Galvis A. [Incidence of yellow fever in different zones of Colombia]. Boletin Oficina Sanit Panam Pan Am Sanit Bur. 1958; 44(1):10–19.

17. Recommendations for Laboratory Detection and Diagnosis of Arbovirus Infections in the Region of the Americas [Internet]. PAHO; 2023 [cited 2026 Aug 19]. Available from: https://iris.paho.org/handle/10665.2/57555

18. Thoisy B de, Silva NIO, Sacchetto L, Trindade G de S, Drumond BP. Spatial epidemiology of yellow fever: Identification of determinants of the 2016-2018 epidemics and at-risk areas in Brazil. PLoS Negl Trop Dis. Public Library of Science; 2020; 14(10):e0008691.

19. Hamrick PN, Aldighieri S, Machado G, et al. Geographic patterns and environmental factors associated with human yellow fever presence in the Americas. PLoS Negl Trop Dis. 2017; 11(9):e0005897.

20. Judson SD, Kenu E, Fuller T, et al. Yellow fever in Ghana: Predicting emergence and ecology from historical outbreaks. PLOS Glob Public Health. Public Library of Science; 2024; 4(10):e0003337.

21. The IUCN Red List of Threatened Species [Internet]. IUCN Red List Threat. Species. [cited 2025 Nov 24]. Available from: https://www.iucnredlist.org/en

22. Johansson MA, Vasconcelos PFC, Staples JE. The whole iceberg: estimating the incidence of yellow fever virus infection from the number of severe cases. Trans R Soc Trop Med Hyg. 2014; 108(8):482–487.

23. Palma AC, Vélez A, Gómez-Posada C, López H, Zárate DA, Stevenson PR. Use of space, activity patterns, and foraging behavior of red howler monkeys (Alouatta seniculus) in an Andean forest fragment in Colombia. Am J Primatol. 2011; 73(10):1062–1071.

24. Bates M, Roca-Garcia M. The Douroucouli (Aotus) in Laboratory Cycles of Yellow Fever. 1945 [cited 2025 Dec 19]; . Available from: https://www.ajtmh.org/view/journals/tpmd/s1-25/5/article-p385.xml

25. Bustamante-Manrique S, Botero-Henao N, Castaño JH, Link A. Activity budget, home range and diet of the Colombian night monkey (Aotus lemurinus) in peri-urban forest fragments. Primates J Primatol. 2021; 62(3):529–536.

26. Sanchez-Rojas IC, Bonilla-Aldana DK, Solarte-Jimenez CL, et al. Fatal yellow fever among captive non-human primates in southern Colombia, 2025. Front Vet Sci [Internet]. Frontiers; 2025 [cited 2025 Nov 21]; 12. Available from: https://www.frontiersin.org/journals/veterinary-science/articles/10.3389/fvets.2025.1655474/full

27. Bonilla-Aldana DK, Bonilla-Aldana JL, Castellanos JE, Rodriguez-Morales AJ. Importance of Epizootic Surveillance in the Epidemiology of Yellow Fever in South America. Curr Trop Med Rep. 2025; 12(1):16.

28. Wilk-da-Silva R, Prist PR, Medeiros-Sousa AR, Laporta GZ, Mucci LF, Marrelli MT. The role of forest fragmentation in yellow fever virus dispersal. Acta Trop. 2023; 245:106983.

