## Appendix for "Yellow fever re-emergence in Tolima, Colombia 2024-25: an eco-epidemiological study"

**Table of Contents**

**1. Review of Historical Outbreaks in Tolima**

*Fig. S1. Historical and recent yellow fever cases by municipality in Tolima, 1830-2025*

**2. Spatial Statistics: Moran’s I**

*Fig. S2. Local Moran’s I map of municipalities in Tolima, Colombia 2024-2025*

*Fig. S3. Local Moran’s I map of veredas in Tolima, Colombia 2024-2025*

**3. Spatial Statistics: Cluster Analysis**

*Fig. S4. Map of reported YF in NHPs in Tolima Colombia*

*Fig. S5. L function non-human primate yellow fever cases*

*Fig. S6. L function of human yellow fever cases*

*Fig. S7. Cross L function, humans vs non-human primate yellow fever cases*

**4. Spatial Data Preparation**

*Fig. S8. Conceptual framework for yellow fever risk in Tolima, Colombia*

*Table S1. Ecological and Sociodemographic Covariates*

*Table S2. Covariate Extraction and Preparation*

*Fig. S9. Map of non-human primate species ranges in Tolima, Colombia.*

**5. Exploratory covariate comparison (veredas with human yellow fever cases vs without)**

*Fig. S10. Overall mean monthly precipitation in veredas with cases vs without*

*Fig. S11. Mean monthly precipitation in veredas with cases vs without*

*Fig. S12. Precipitation anomaly during outbreak period vs historical periods*

*Fig. S13. Overall mean monthly temperature in veredas with cases vs without*

*Fig. S14. Mean monthly temperature in veredas with cases vs without*

*Fig. S15. Temperature anomaly during outbreak period vs historical periods*

*Table S3. Summary of elevation at case locations*

*Fig. S16. Mean elevation in veredas with cases vs those without*

*Table S4. Proportion of land cover at case locations*

*Fig. S17. Map of landcover in Tolima, Colombia*

*Fig. S18. Mean Normalized Difference Vegetation Index (NDVI) in veredas with cases vs those without*

*Fig. S19. Mean population density in veredas with cases vs those without*

*Fig. S20. Global Gridded Relative Deprivation Index (GRDI) in veredas with cases vs those without*

*Fig. S21. Mean built-up surface in veredas with cases vs those without*

**6. Logistic Regression Analysis of Mortality**

*Table S5. Summary of Model Results*

**7. Bayesian Hierarchical Spatiotemporal Model**

*Table S6. Summary of Model Results and Diagnostics*

*Table S7. Percentage of Random-Effect Variance Explained by Model Component*

**1. Review of Historical Outbreaks in Tolima**

To identify historical yellow fever outbreaks in Colombia, we searched across several electronic databases, including PubMed/MEDLINE, the Directory of Open Access Journals, and the Natural Science Collection, amongst others, using the University of California Library. Search terms included “yellow fever” AND “Colombia,” which were also used to search Google Scholar. The search was conducted between October 2^nd^, 2025 through December 9^th^, 2025.

Yellow fever cases were reported in Tolima in 1830 (Ambalema, Honda), 1866 (Espinal, Venadillo), 1880 (Purificación), and 1940 (Mariquita). In 1958, Dr. Augusto Gash Galvis published a study on yellow fever incidence based on post-mortem human liver samples via viscerotomy, corresponding to the period 1934–1956. This study reported positive samples for yellow fever in the department of Tolima, in the municipalities of Armero (n = 1) and Mariquita (n = 10).

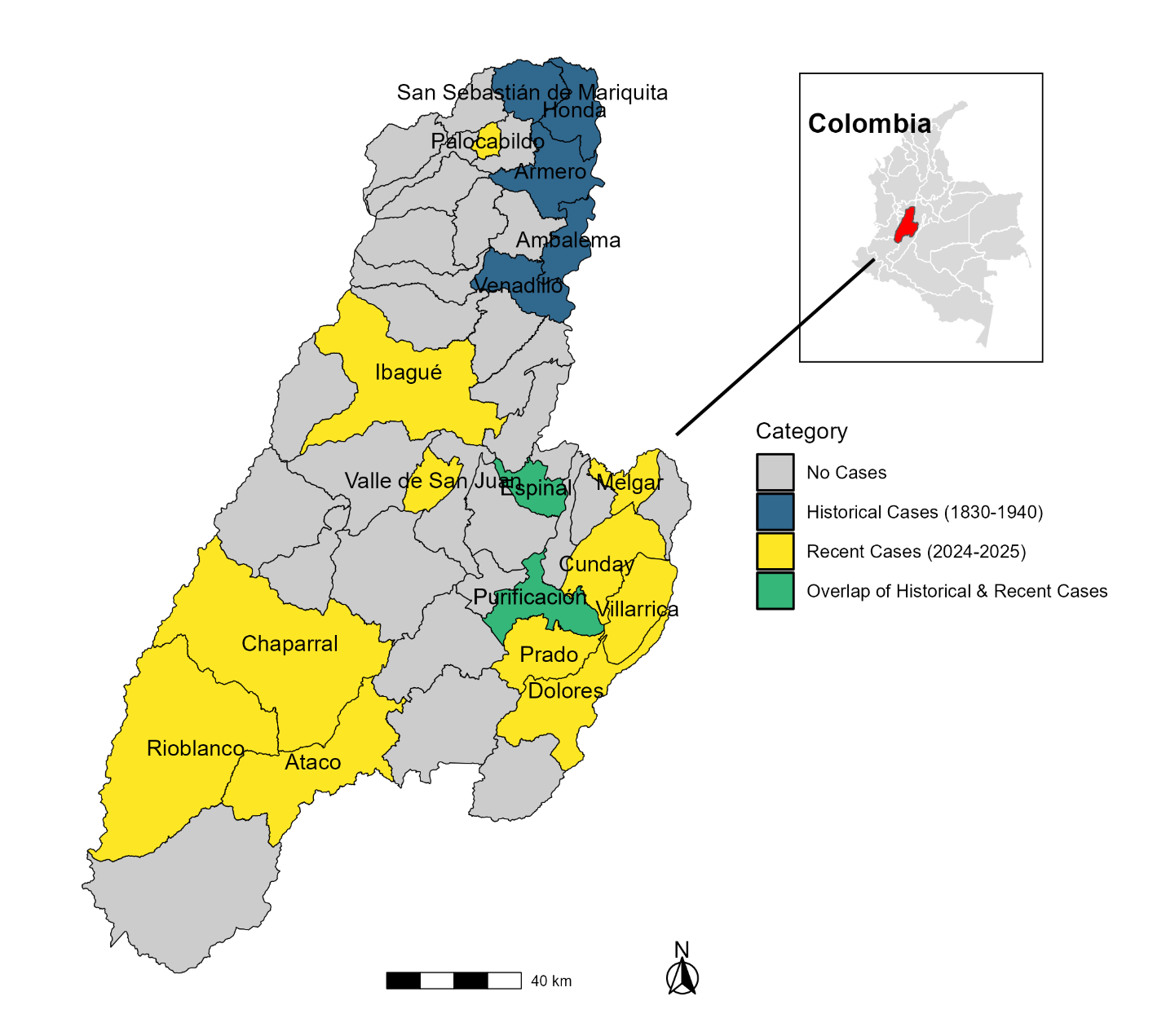

**Fig. S1. Historical and recent yellow fever cases by municipality in Tolima, 1830-2025**

The map depicts the municipalities of reported historical yellow fever cases (1830-1940) as well as cases during the outbreak study period (2024-2025).

**2. Spatial Statistics: Moran’s I**

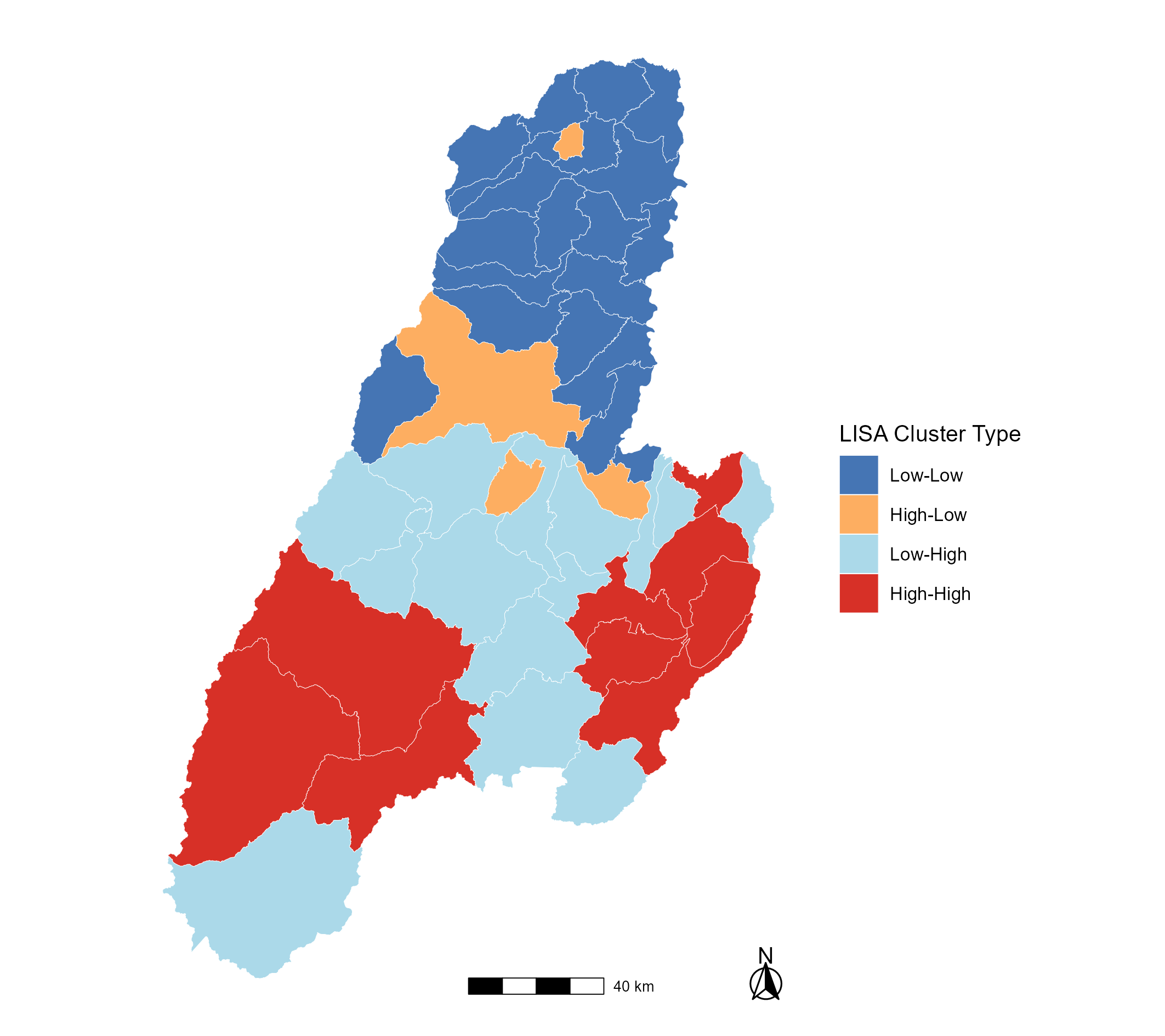

**Fig. S2. Local Moran’s I map of municipalities in Tolima, Colombia 2024-2025**

The map displays Local Indicators of Spatial Association (LISA) at the municipality level, calculated via local Moran’s I of yellow fever incidence rates during the 2024-2025 outbreak period. The median-based quadrant classification labels each municipality as high or low depending on whether its incidence rate is above or below the overall median and the high or low status of its neighbors. The resulting cluster types are: High-High (red) denoting areas with above-median incidence rates surrounded by neighbors that are also high, Low-Low (dark blue) areas have below-median incidence rates surrounded by neighbors with low rates, High-Low (orange) areas have above-median values surrounded by neighbors with low rates, and Low-High (light blue) areas have below-median rates and are surrounded by neighbors with high rates. Municipality boundaries are shown in light grey.

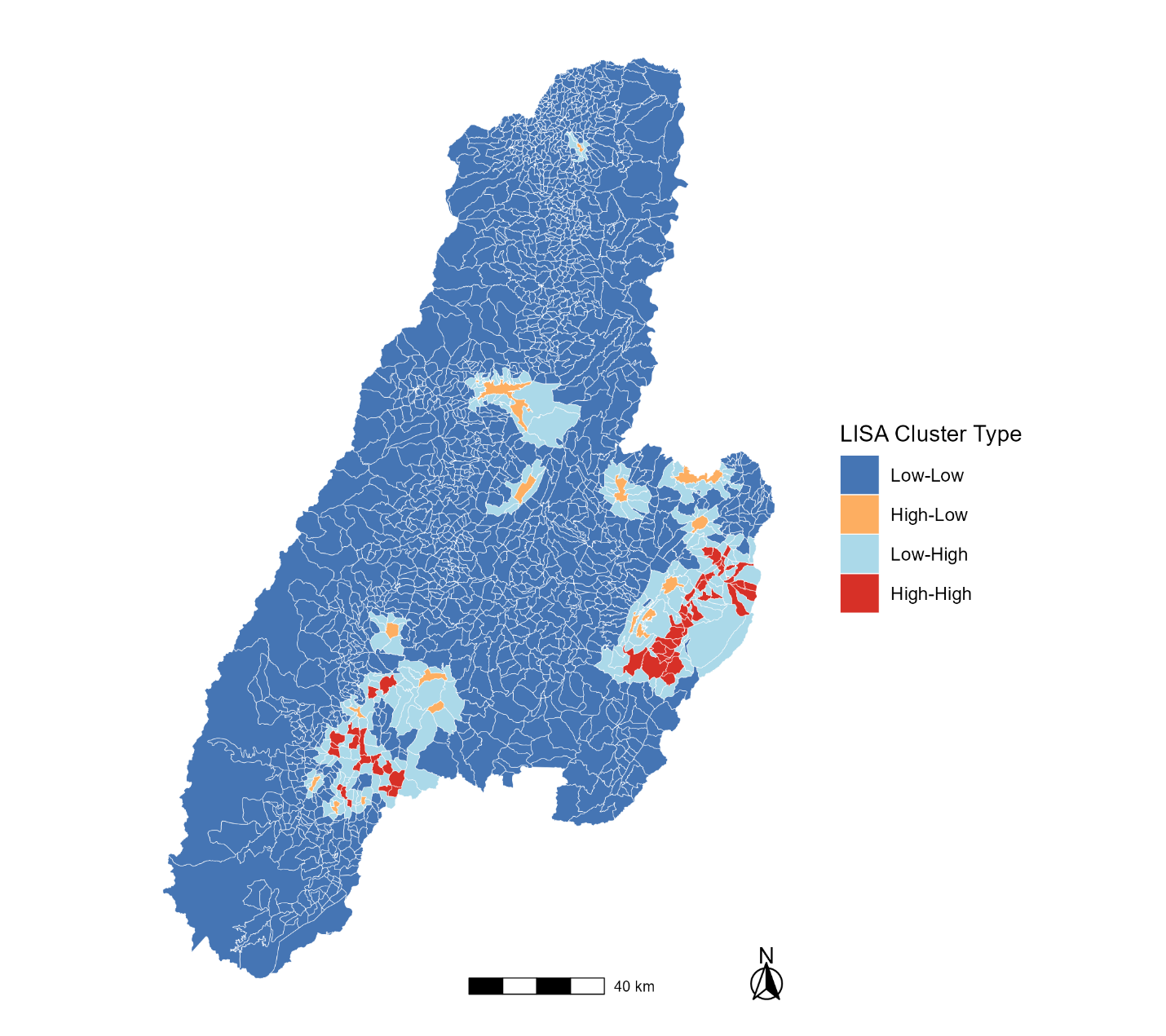

**Fig. S3. Local Moran’s I map of veredas in Tolima, Colombia 2024-2025**

The map displays Local Indicators of Spatial Association (LISA) at the vereda level, calculated via local Moran’s I of yellow fever incidence rates during the 2024-2025 outbreak period. The median-based quadrant classification labels each municipality as high or low depending on whether its incidence rate is above or below the overall median and the high or low status of its neighbors. The resulting cluster types are: High-High (red) denoting areas with above-median incidence rates surrounded by neighbors that are also high, Low-Low (dark blue) areas have below-median incidence rates surrounded by neighbors with low rate, High-Low (orange) areas have above-median incidence rates surrounded by neighbors with low rates, and Low-High (light blue) areas have below-median rates and are surrounded by neighbors with high rates. Vereda boundaries are shown in light gray.

**3. Spatial Statistics: Cluster Analysis**

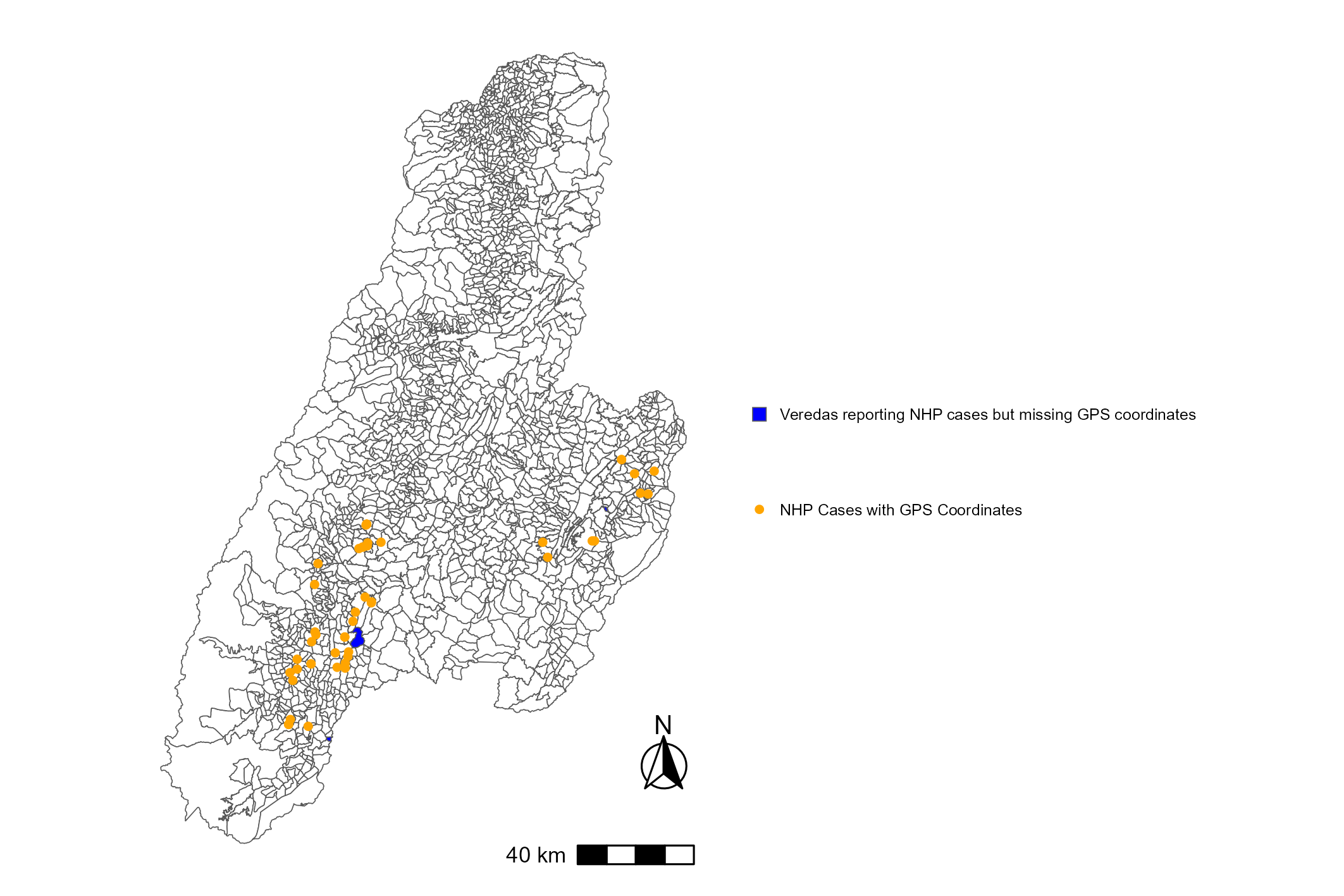

**Fig S4. Map of reported YF in NHPs in Tolima, Colombia 2024-2025**

This map displays YF cases in NHPs reported to the Tolima Department of Public Health from February 2024 – October 2025. GPS coordinates were provided for 49 of the 53 reported cases and are mapped as orange points. Veredas that reported NHP cases without accompanying GPS coordinates are shown in blue, including Copete Oriente (1 case), El Diviso (2 cases), and San Isidro in the Cunday municipality (1 case).

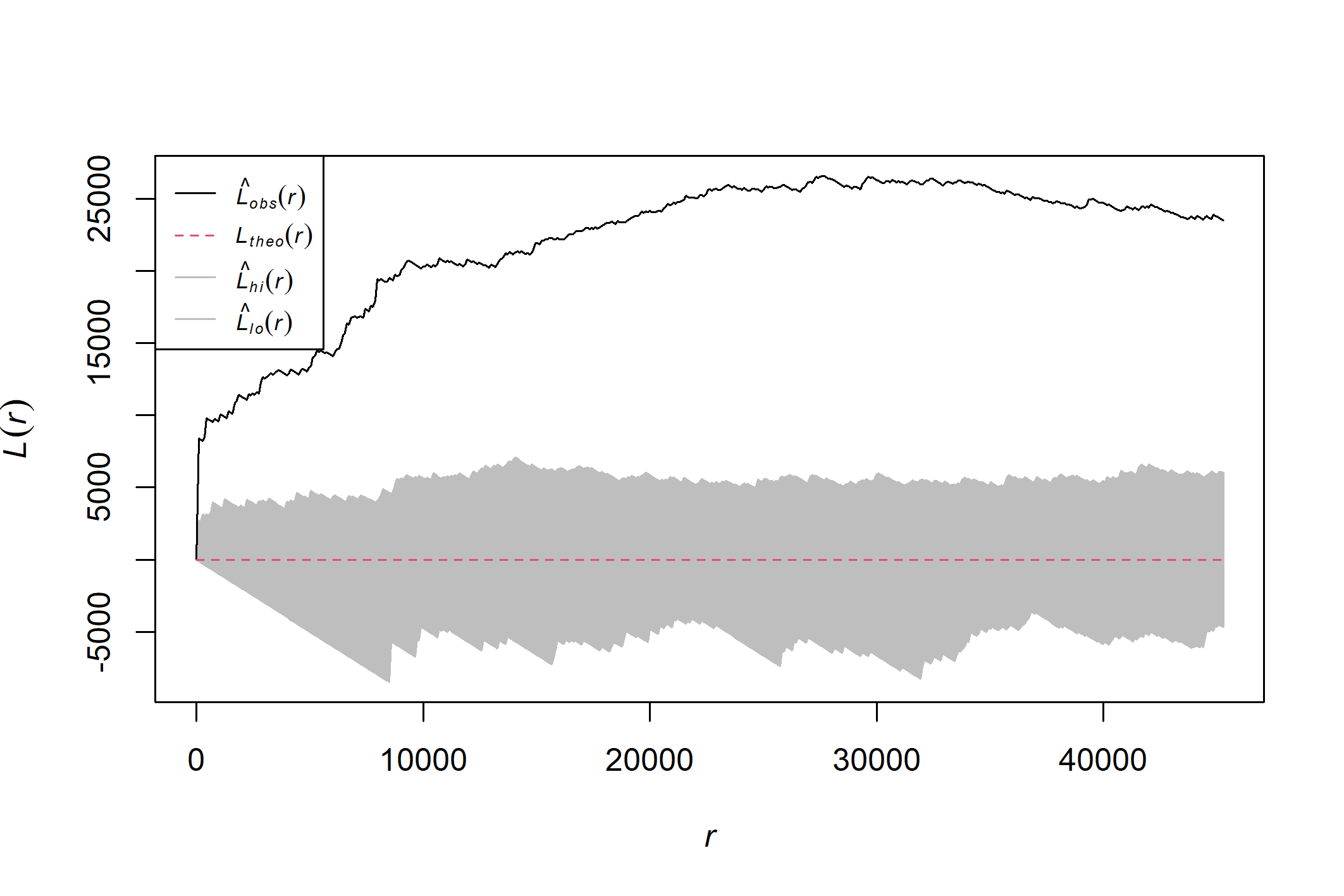

**Fig. S5. L function non-human primate yellow fever cases**

The plot displays Ripley’s L-r function with a 95% Monte Carlo envelope calculated using 999 simulations and an isotropic correction for non-human primate yellow fever cases (n =49) in Tolima, Colombia. Non-human primate cases without geographic coordinates were excluded from this analysis (n=4). L_obs_ represents the observed L(r) – r function at distance r while L_theo_ represents the theoretical L(r) – r function under complete spatial randomness. L_hi_ and L_lo_ represent the upper and lower confidence envelopes around complete spatial randomness, respectively. Positive deviations of Lobs above L_theo_ indicates spatial clustering at distance, r with higher values of L-r indicating stronger clustering. Clustering was strongest at a distance of 27.6 km.

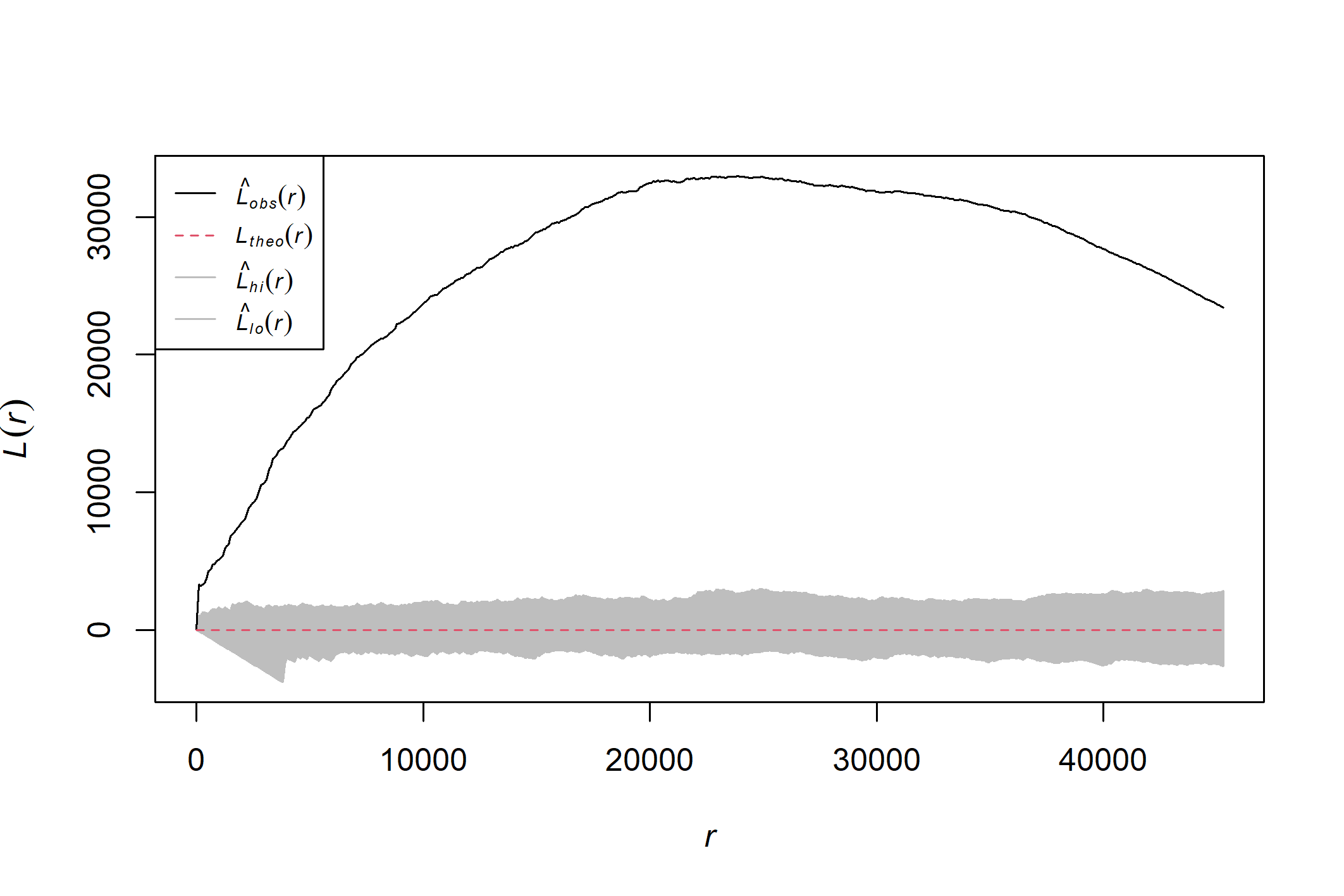

**Fig. S6. L function of human yellow fever cases**

The plot displays Ripley’s L-r function with a 95% Monte Carlo envelope calculated using 999 simulations and an isotropic correction for human yellow fever cases in Tolima, Colombia (n=116). L_obs_ represents the observed L(r) – r function at distance r while L_theo_ represents the theoretical L(r) – r function under complete spatial randomness. L_hi_ and L_lo_ represent the upper and lower confidence envelopes around complete spatial randomness, respectively. Positive deviations of L_obs_ above L_theo_ indicates spatial clustering at distance, r with higher values of L-r indicating stronger clustering. Clustering was strongest at a distance of 23.9 km.

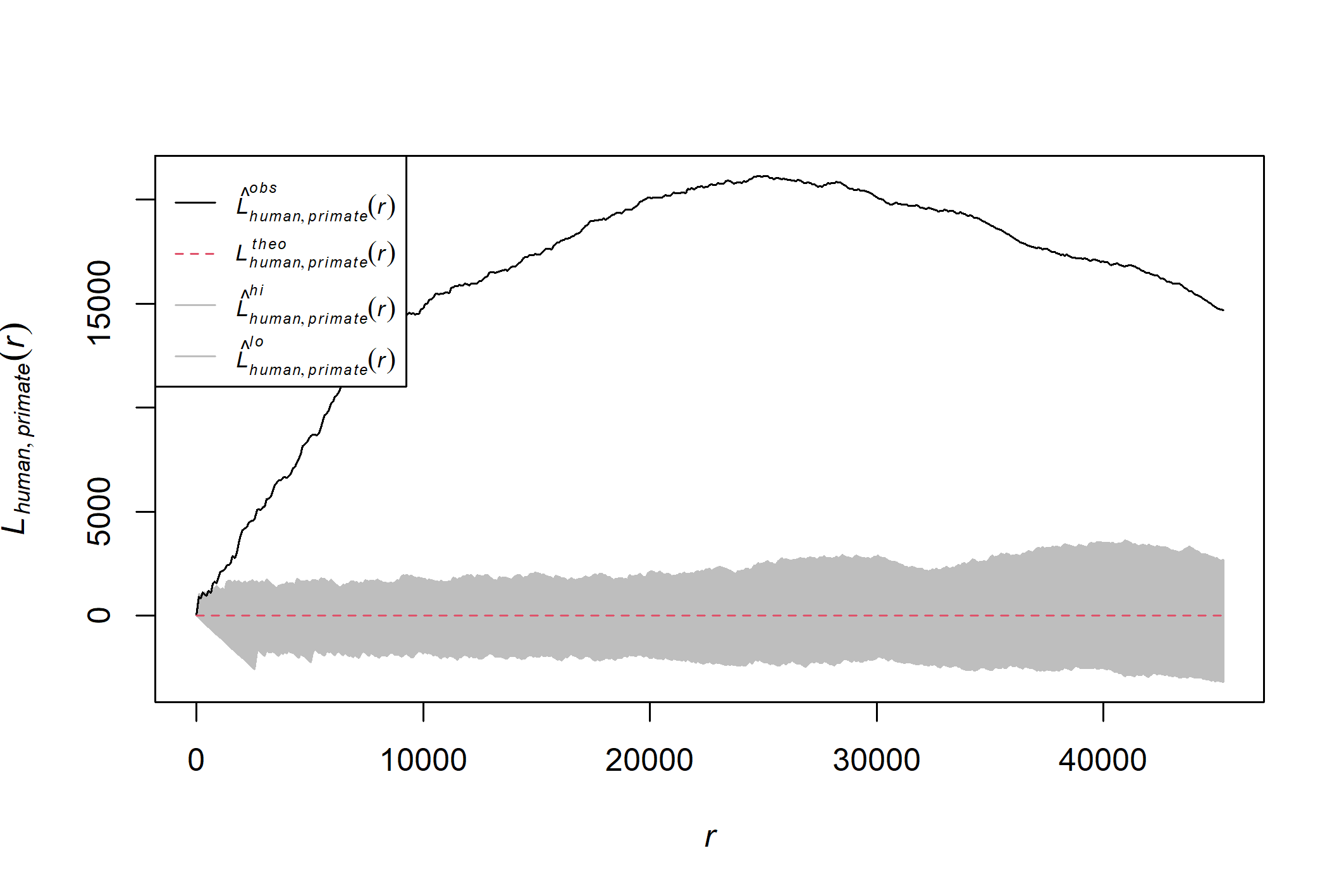

**Fig. S7. Cross L function, humans vs non-human primate yellow fever cases**

The plot displays Ripley’s cross L-r function with a 95% Monte Carlo envelope calculated using 999 simulations and an isotropic correction for human and non-human primate yellow fever cases in Tolima, Colombia. L_obs_ represents the observed cross L(r) – r function at distance r while L_theo_ represents the theoretical L(r) – r function under the null hypothesis of spatial independence between human and non-human primate cases. L_hi_ and L_lo_ represent the upper and lower confidence envelopes, respectively. Positive deviations of Lobs above L_theo_ indicates spatial clustering between human and non-human primate cases at distance, r with higher values of L-r indicating stronger clustering. Clustering was strongest at a distance of 24.9 km.

**4. Spatial Data Preparation**

Shapefiles for municipalities and veredas were obtained via Humanitarian Data Exchange under a CC BY-IGO license (https://data.humdata.org/dataset/cod-ab-col). Spatial autocorrelation was assessed using global and local versions of Moran’s I, calculated using the spdep package in R. The incidence of YF cases was calculated using WorldPop data aggregated to the municipality and vereda levels.

We used the conceptual framework below to identify key covariates for the spatial analysis. For our exploratory analysis and modeling, we extracted the mean values for each covariate at the vereda level. We also extracted elevation and land-cover values at specific residence locations. For modeling, a correlation matrix was used to exclude covariates that had a correlation coefficient of >|0.7|. There was a strong correlation between temperature and elevation (-0.8) and poverty/deprivation and built-up surface (-0.7), so we kept temperature and poverty/deprivation in the model based on our conceptual framework.

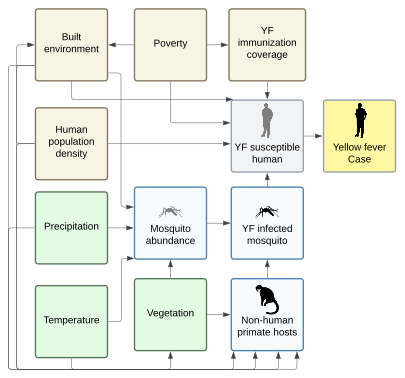

**Fig. S8. Conceptual framework yellow fever risk in Tolima, Colombia**

The figure depicts key ecological (green) and sociodemographic (brown) variables considered in relation to human yellow fever cases in Tolima. This conceptual framework was used to identify which covariates to include in exploratory spatial analysis and inferential spatiotemporal modeling.

Veredas are the smallest rural administrative divisions within a municipality in Colombia. While many veredas are officially recognized by the Departamento Administrativo Nacional de Estadístisca (DANE), others are recognized informally at the local level, leading to discrepancies in vereda names and boundaries. We used the shapefile of veredas based on the DANE classification obtained from Humanitarian Data Exchange. The original shapefile contained 47 unassigned areas, including small gaps in polygons and polygons containing high population areas like the capital city of Ibagué. To accurately extract zonal statistics for covariates, unassigned areas in the vereda shapefile were corrected in R using the sf package. The polygons of the vereda shapefile were first unioned and then subtracted from the municipality shapefile to identify unassigned gaps. Gaps were assigned to the geographically nearest vereda using the st_nearest_feature and merged with the vereda geometry to produce a complete shapefile. The average size of veredas in the final shapefile was 12.22km (range: 0.18 – 1107.7km).

For YF cases among humans and NHPs with GPS coordinates available, we used these coordinates to assign cases to veredas in the updated shapefile using the st_join and st_intersects functions from the R sf package. The assigned veredas were then compared with locally recognized vereda names used by the Secretaria de Salud del Tolima. The complete list of vereda names used in the analysis, along with the corresponding names from the Secretaria de Salud del Tolima, and R markdown files for the analysis are available online at: <https://doi.org/10.6084/m9.figshare.31061575>

| **Table S1. Ecological and Sociodemographic Covariates** | | | |
| --- | --- | --- | --- |
| Covariate | Details | Resolution | Source |
| Monthly mean precipitation  2024-2025 | Satellite-based thermal infrared rainfall estimates with in-situ station observations | 0.05° | CHIRPS v3 |
| Monthly mean temperature  2024-2025 | Satellite-based land surface temperature | 0.05° | MODIS/061/MOD21C3 |
| Elevation | Derived from the Shuttle Radar Topography (SRTM) elevation data | 30 arc seconds/1 km^2^ | Worldclim 2.1 |
| Monthly Vegetation (NDVI)  2024-2025 | Normalized Difference Vegetation Index | 1 km^2^ | MODIS/061/MOD13A3 |
| Annual Landcover  2024 | Satellite-based with machine learning landcover classification | 30 m^2^ | MapBiomas |
| Population density  2025 | Gridded population estimates | 100 m^2^ | Worldpop |
| Deprivation and Poverty  2020 | Weight indices of sociodemographic and satellite data | 1 km^2^ | Global Gridded Relative Deprivation Index, Version 1 |
| Built-up surface area 2025 | Built-up surface based on Sentinel Earth Observation data | 100 m^2^ | Copernicus Global Human Settlement Layer |

**Table S2. Covariate Extraction and Preparation**

| **Covariate** | **Export Source** | **Extraction Method** |
| --- | --- | --- |
| Monthly mean precipitation  2024-2025 | **Descriptive comparisons between veredas with and without cases:**  Mean monthly precipitation values from June 2024-October 2025 were saved as GeoTIFF files from the CHIRPS v3 monthly datasets in the Latin America region (<https://data.chc.ucsb.edu/products/CHIRPS/v3.0/monthly/latam/tifs/>). | Raster files were imported into R and cropped to the study area using the vereda shapefile using the terra package. For boxplot visualization, average monthly precipitation values were summarized across the study period (September 2024–October 2025), and month-specific mean temperatures were also extracted for each vereda using the exact_extract function. |
|  | **Modeling:**  Mean monthly precipitation values from June 2024-October 2025 were saved as GeoTIFF files from the CHIRPS v3 monthly datasets in the Latin America region (<https://data.chc.ucsb.edu/products/CHIRPS/v3.0/monthly/latam/tifs/>). | The mean precipitation value for each month was assigned to each vereda polygon by the polygon’s unique object ID. |
| Monthly mean temperature  2024-2025 | **Descriptive comparisons between veredas with and without cases:**  The average monthly land surface daytime temperature (LST) and the average LST for each month from September 2024-October 2025 was extracted for each vereda using Google Earth Engine. Vereda boundaries were uploaded as a shapefile, and temperature values were obtained from the MODIS/061/MOD21C3 instrument. Data was exported as a GeoTIFF file. | Raster files were imported into R using the terra package. For boxplot visualization, average monthly temperature values were summarized across the study period (September 2024–October 2025), and month-specific mean temperatures were also extracted for each vereda.  86 missing month-specific mean temperature values were imputed using the mean temperature of neighboring veredas in R. Neighbors were defined using queen contiguity. |
|  | **Modeling:**  Monthly mean LST values were extracted for each vereda using Google Earth Engine. Vereda boundaries were uploaded as a shapefile, and monthly mean temperature values were obtained from the MODIS/061/MOD21C3 instrument for the period June 2024 – October 2025. Data was exported as a CSV file. | The mean temperature value for each month was assigned to each vereda polygon by the polygon’s unique object ID. 86 missing monthly mean temperature values were imputed using the mean temperature of neighboring veredas in R. Neighbors were defined using queen contiguity. |
| Elevation | An elevation GeoTIFF file for Colombia was exported from WorldClim. | The raster file was imported into R using the terra package. To create boxplots of average elevation, a mean elevation for each vereda was extracted using the exact_extract function. To summarize elevation at case locations, the elevation at each case’s coordinates was extracted using the extract function in the terra package and the minimum, maximum, mean, and standard deviation of elevation were calculated. |
| Monthly Vegetation (NDVI)  2024-2025 | **Descriptive comparisons between veredas with and without cases:**  The average NDVI from September 2024-October 2025 was extracted for each vereda using Google Earth Engine. Vereda boundaries were uploaded as a shapefile, and NDVI values were obtained from the MODIS/061/MOD13A3 instrument. Data was exported as a GeoTIFF file. | The raster file was imported into R and cropped to the study area using the vereda shapefile using the terra package. The exact_extract function was used to assign monthly mean NDVI values to each vereda polygon. |
|  | **Modeling:**  Monthly mean NDVI values were extracted for each vereda using Google Earth Engine. Vereda boundaries were uploaded as a shapefile, and monthly mean NDVI values were obtained from the MODIS/061/MOD13A3 instrument for the period June 2024 – October 2025. Data was exported as a CSV file. | The mean NDVI value for each month was assigned to each vereda polygon by the polygon’s unique object ID. |
| Annual Landcover  2024 | A categorical 2024 landcover GeoTIFF file was exported for the department of Tolima from MapBiomas. | The raster was imported into R using the terra package. For areal analysis, the number of raster cells for each land cover class were counted within each vereda using the extract function. The proportion of each land cover class was calculated by dividing the pixel counts for each classification by the total pixels within each vereda.  To summarize land cover classes at case locations, the class each case’s coordinates was extracted using the extract function in the terra package. |
| Population density  2025 | A GeoTIFF of Colombia’s project 2025 population was downloaded from WorldPop. | The raster file was imported into R and cropped to the study area using the vereda shapefile using the terra package. The exact_extract function was used to assign the sum population values for each vereda polygon to calculate its total population. The total area of each polygon was obtained using the st_area function in the sf package, and population density was calculated for each vereda by dividing the total population by the area. |
| Deprivation and Poverty  2020 | GRDI values were obtained from NASA EarthData Search and exported as a GeoTIFF. Data were downloaded for the year 2020 and spatially subset to Colombia using a bounding box covering the study area. | The raster file was imported into R and cropped to the study area using the vereda shapefile using the terra package. The exact_extract function was used to assign mean GRDI values to each vereda polygon. |
| Built-up surface area 2025 | Total built-up surface area for epoch 2025 was obtained from the Copernicus Global Human Settlement Layer, GHS-BUILT-S - R2023A dataset and exported as a GEOTIFF. | The raster file was imported into R and cropped to the study area using the vereda shapefile using the terra package. The exact_extract function was used to assign mean built-up surface area values to each vereda polygon. |

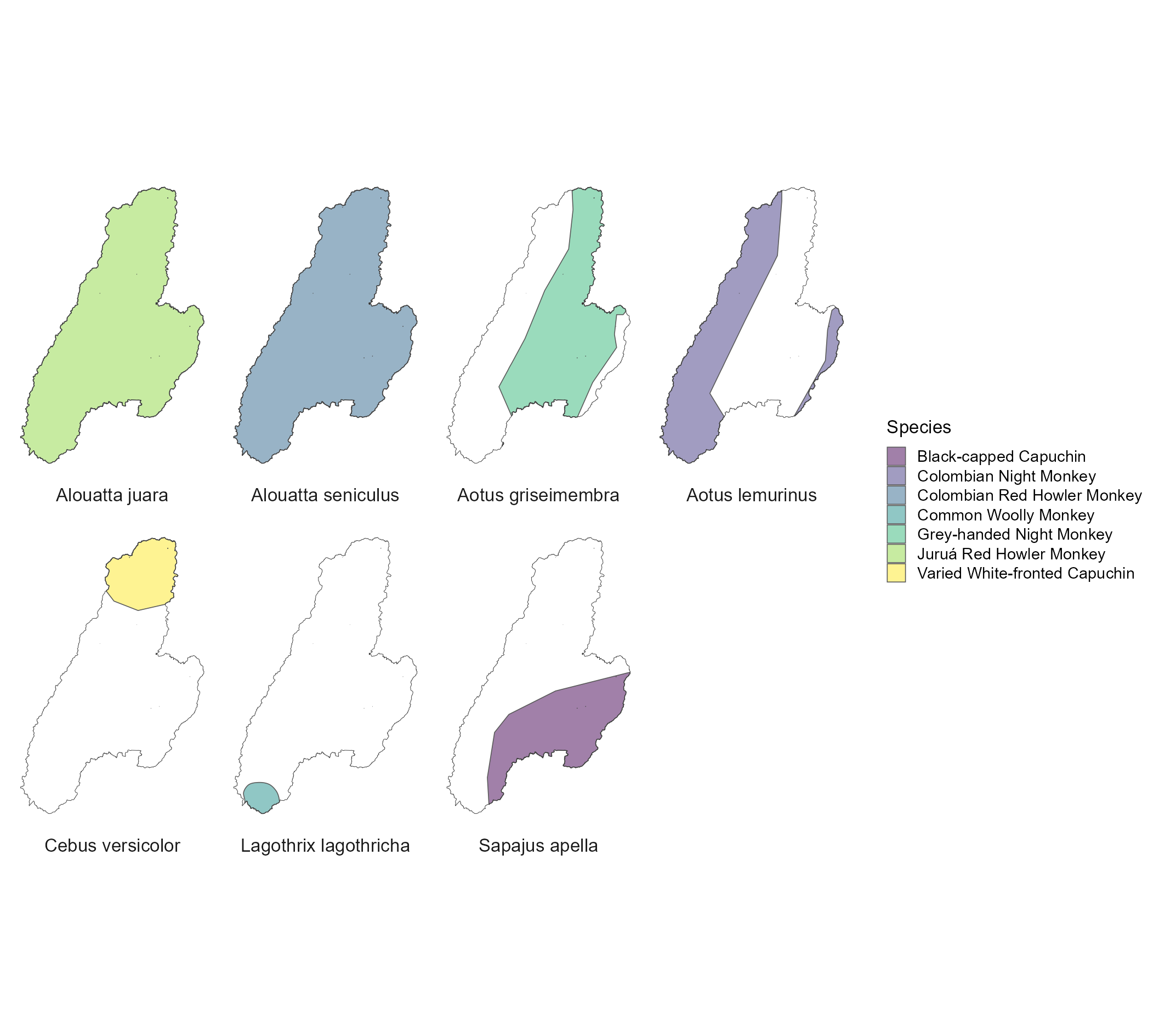

**Fig. S9. Map of non-human primate species ranges in Tolima, Colombia.**

Colored polygons represent the known ranges of each species. The shapefiles for the ranges were obtained from the IUCN Redlist.

**5. Exploratory covariate comparison (veredas with human yellow fever cases vs without)**

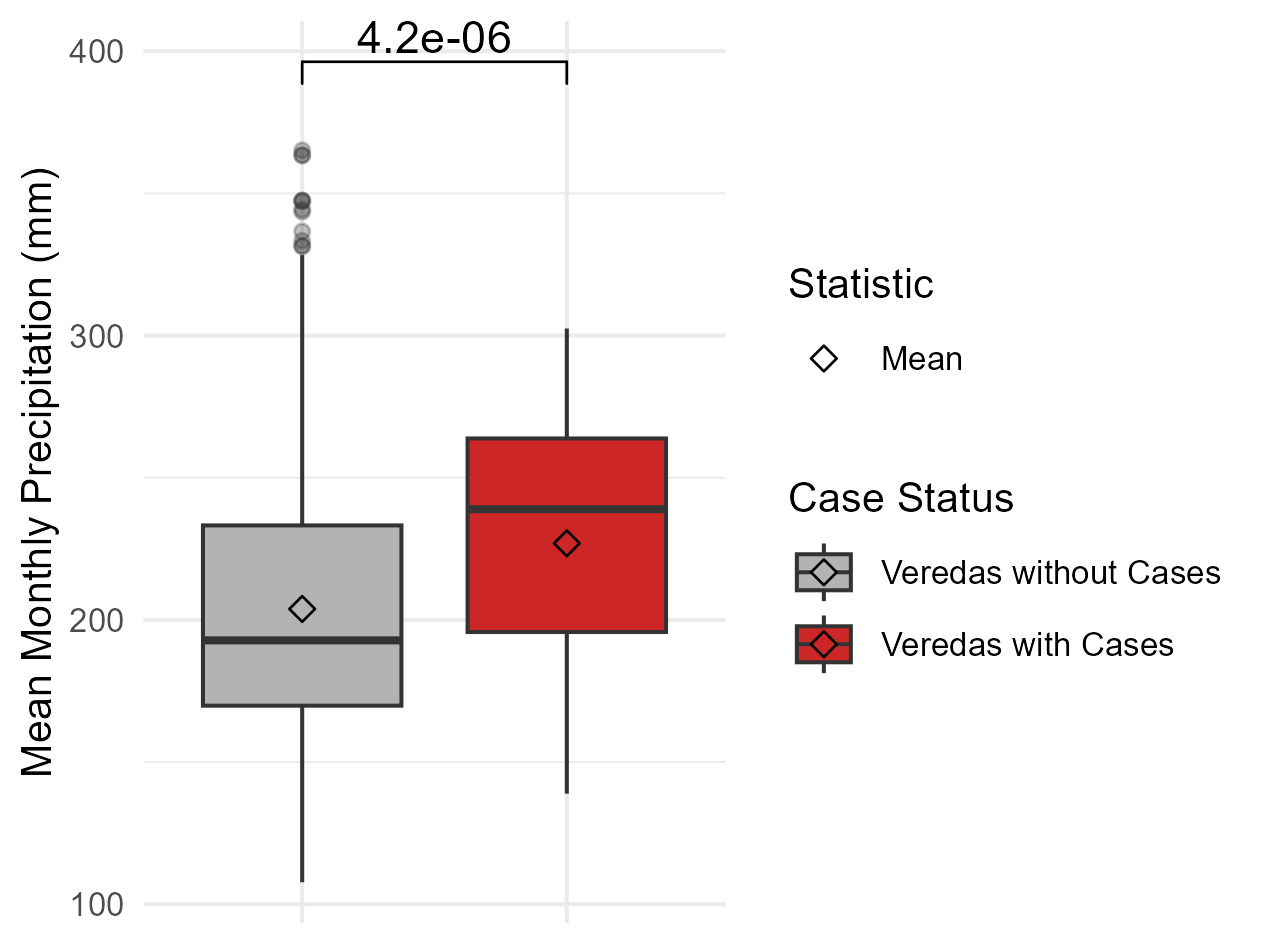

**Fig. S10. Overall mean monthly precipitation in veredas with cases vs without**

Boxplot of average monthly precipitation (mm) in veredas with and without human yellow fever cases in Tolima, Colombia. The central black line in each box represents group median, and the lower and upper limits of the box denote the first and third quartiles, respectively. Diamonds indicate group means. Normality was assessed using a Shapiro-Wilk test, and because precipitation distributions were non-normal, a Wilcoxon rank-sum test was used to determine statistical significance between groups. The resulting p-value is displayed above the boxes. Monthly precipitation data from September 2024 – October 2025 was sourced from CHIRPS v3 at a resolution of 0.05°.

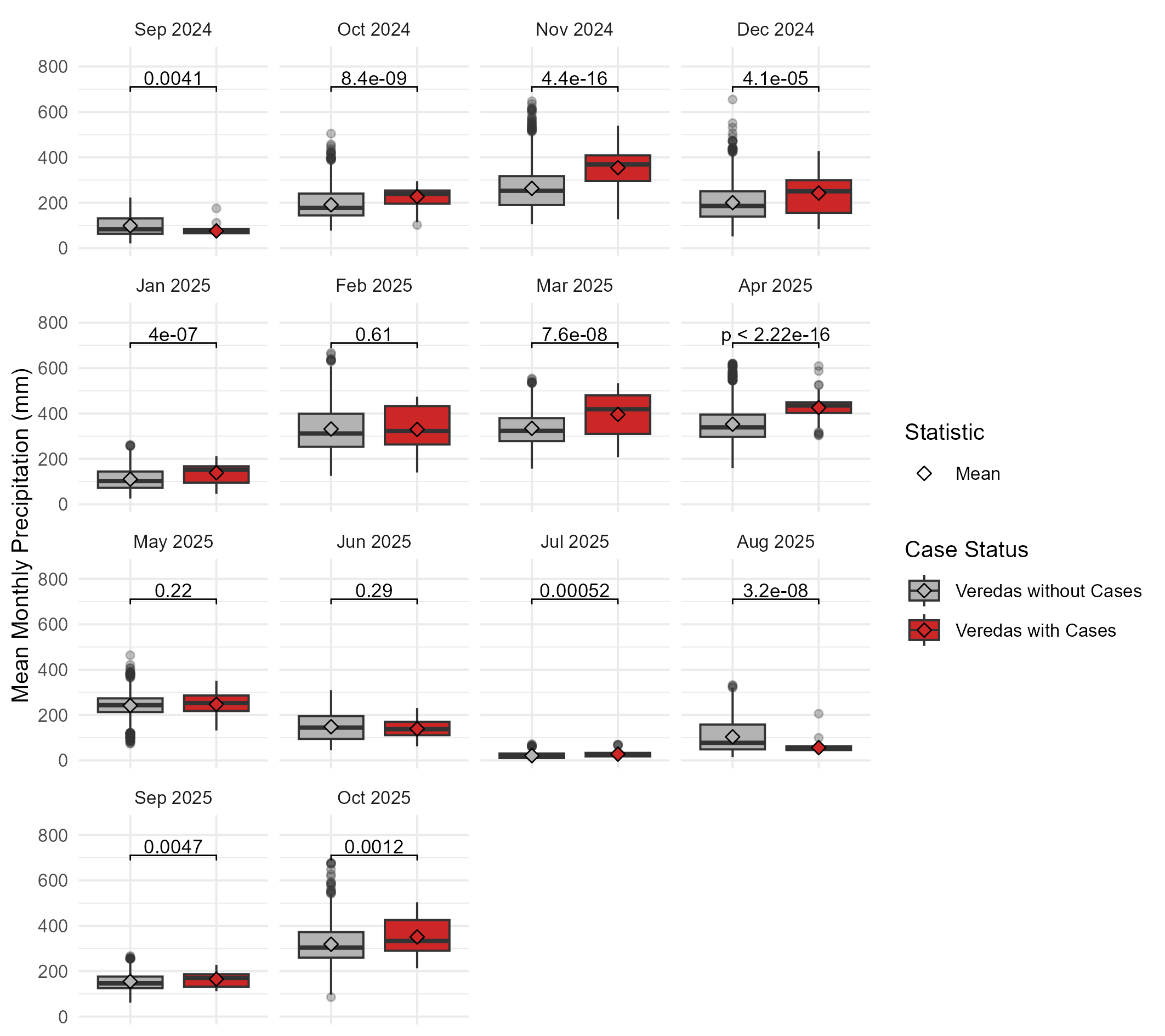

**Fig. S11. Mean monthly precipitation in veredas with cases vs without**

Boxplot of average precipitation (mm) by month in veredas with and without human yellow fever cases in Tolima, Colombia. The monthly mean precipitation was significantly higher in 8 of 12 months in veredas with YF cases from September 2024-2025. The central black line in each box represents group median, and the lower and upper limits of the box denote the first and third quartiles, respectively. Diamonds indicate group means. Normality was assessed using a Shapiro-Wilk test, and because monthly precipitation distributions were non-normal, a Wilcoxon rank-sum test was used to determine statistical significance between groups. The resulting p-value is displayed above the boxes. Monthly precipitation data from September 2024 – October 2025 was sourced from CHIRPS v3 at a resolution of 0.05°.

**(a)**

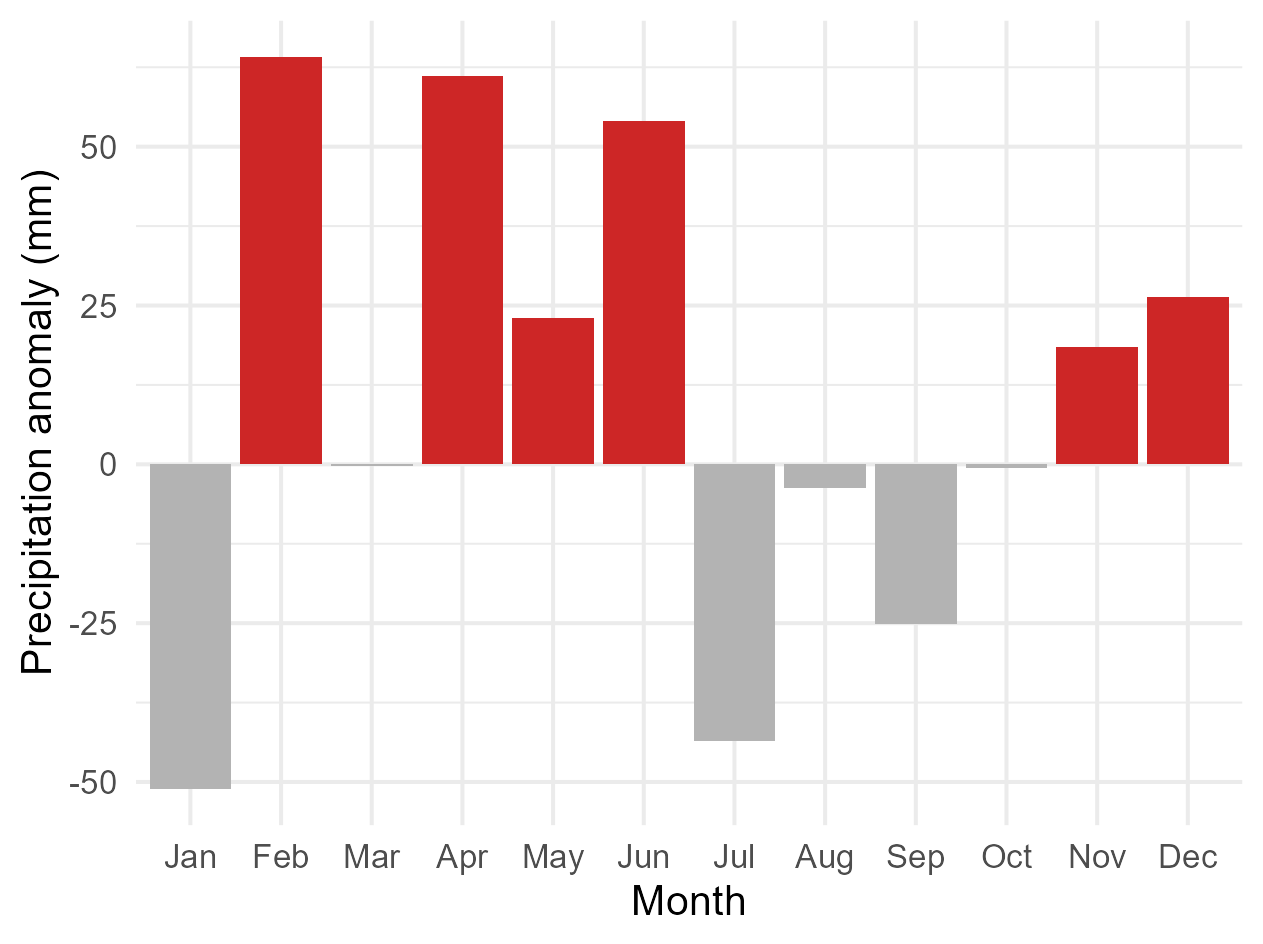

**(b)**

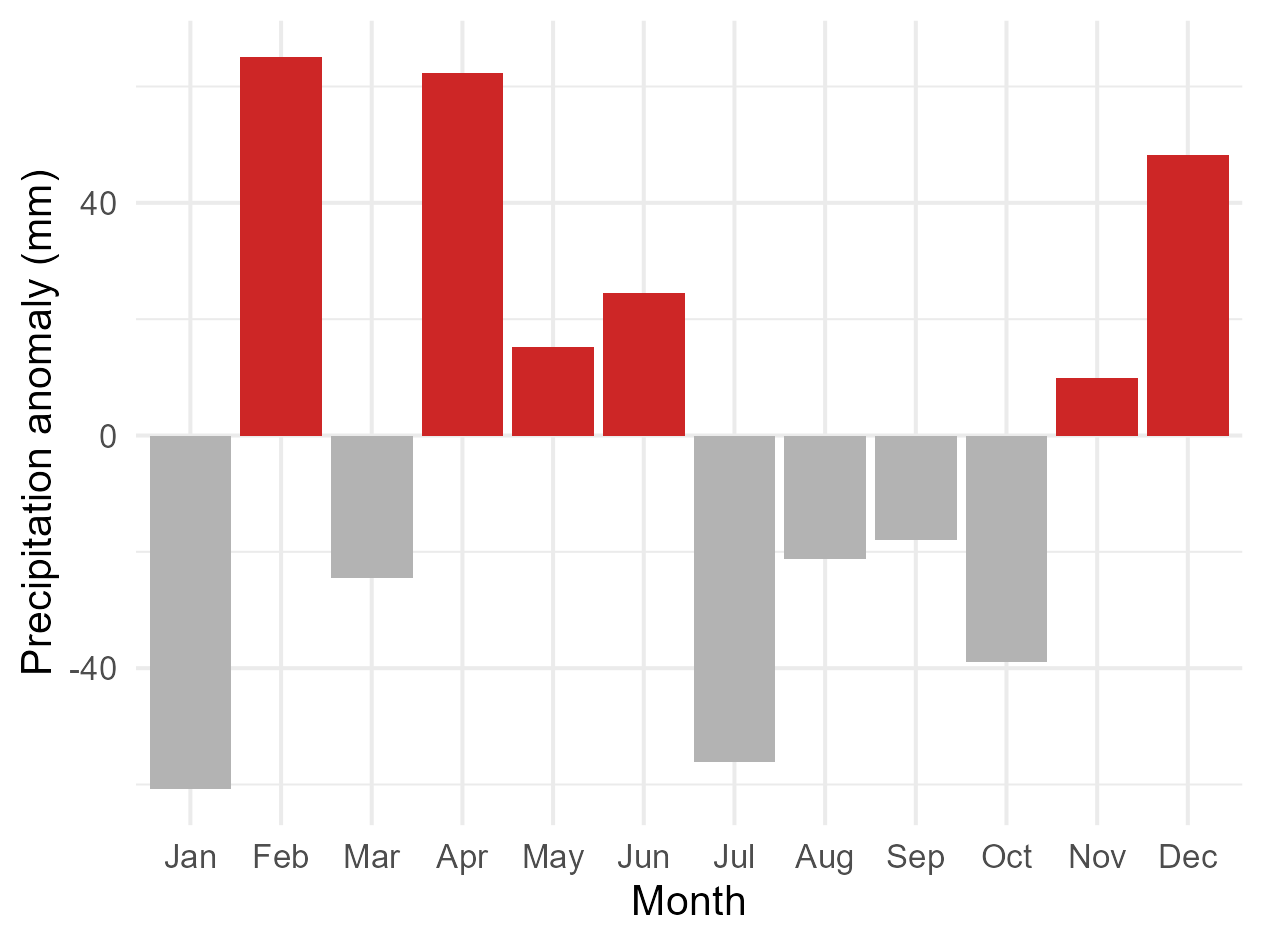

**Fig S12. Precipitation anomaly during outbreak period vs historical periods**

Bar chart of the difference in average precipitation by month between study period (2024-2025) vs (a) 30-year historic period (1993-2023) and (b) and 5-year historic period (2018-2023). Mean monthly precipitation values were first aggregated for each month at the vereda level and then averaged across all veredas in Tolima. The precipitation anomaly for each month was calculated by taking the difference between the study period mean and historical period mean. Bars are colored gray when the study period had lower average monthly precipitation than the historic period and red when average monthly precipitation was higher. Monthly precipitation data from January 1991 – October 2025 was sourced from CHIRPS v3 at a resolution of 0.05°.

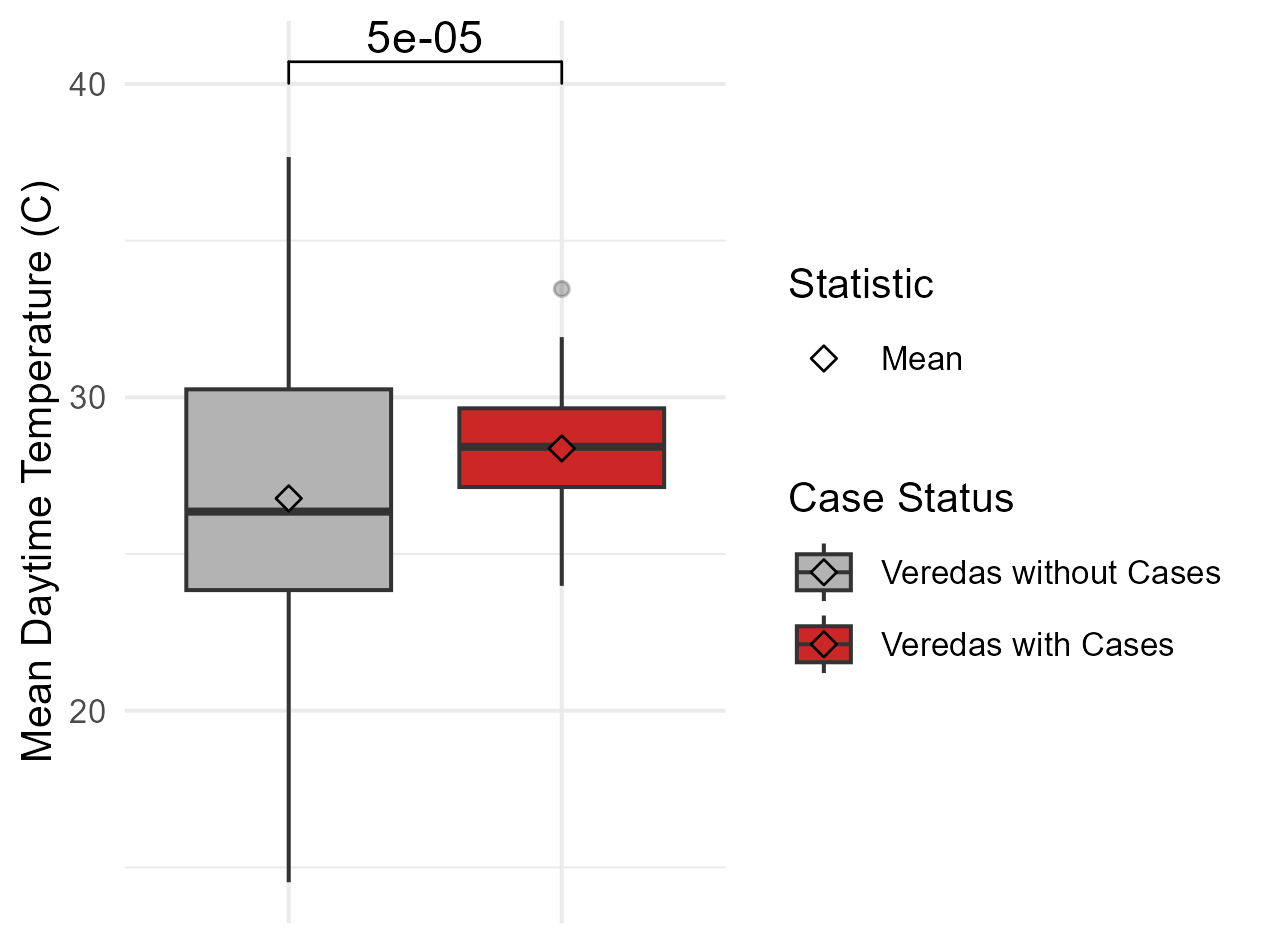

**Fig S13. Overall mean monthly temperature in veredas with cases vs without**

Boxplot of average daytime monthly temperature (℃) in veredas with and without human yellow fever cases in Tolima, Colombia. The central black line in each box represents group median, and the lower and upper limits of the box denote the first and third quartiles, respectively. Diamonds indicate group means. Normality was assessed using a Shapiro-Wilk test, and because temperature distributions were non-normal, a Wilcoxon rank-sum test was used to determine statistical significance between groups. The resulting p-value is displayed above the boxes. Monthly land surface temperature from September 2024 – October 2025 was sourced from MODIS/061/MOD21C3 at a resolution of 0.05°.

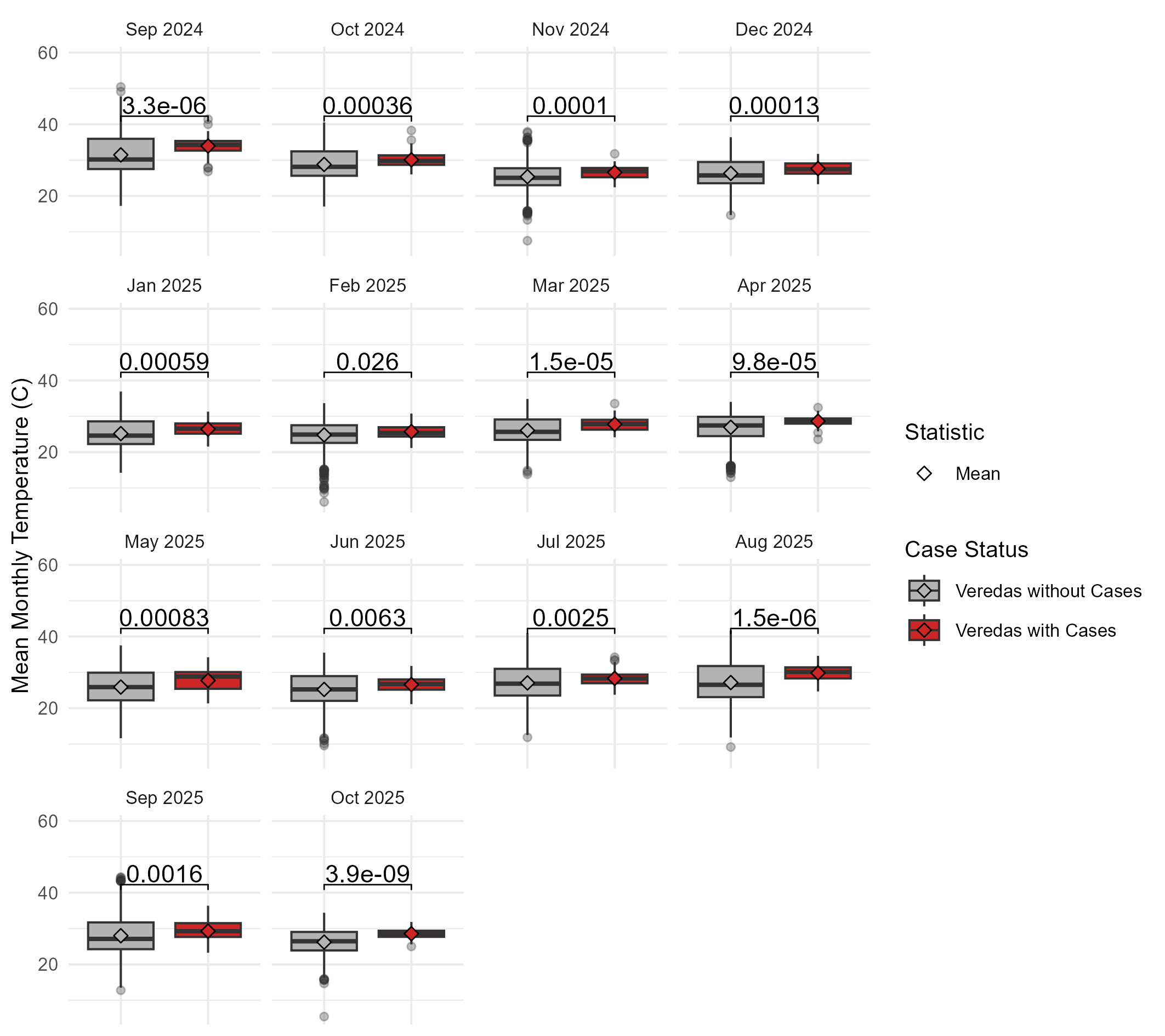

**Fig S14. Mean monthly temperature in veredas with cases vs without**

Boxplot of average daytime temperature (℃) by month in veredas with and without human yellow fever cases in Tolima, Colombia. The monthly mean temperature was significantly higher in 12 of 12 months in veredas with YF cases from September 2024 to 2025. Eighty-one missing values were imputed using the means from spatial neighbors defined by queen contiguity. The central black line in each box represents group median, and the lower and upper limits of the box denote the first and third quartiles, respectively. Diamonds indicate group means. Normality was assessed using a Shapiro-Wilk test, and because monthly temperature distributions were non-normal, a Wilcoxon rank-sum test was used to determine statistical significance between groups. The resulting p-value is displayed above the boxes. Monthly land surface temperature from September 2024 – October 2025 was sourced from MODIS/061/MOD21C3 at a resolution of 0.05°.

**(a)**

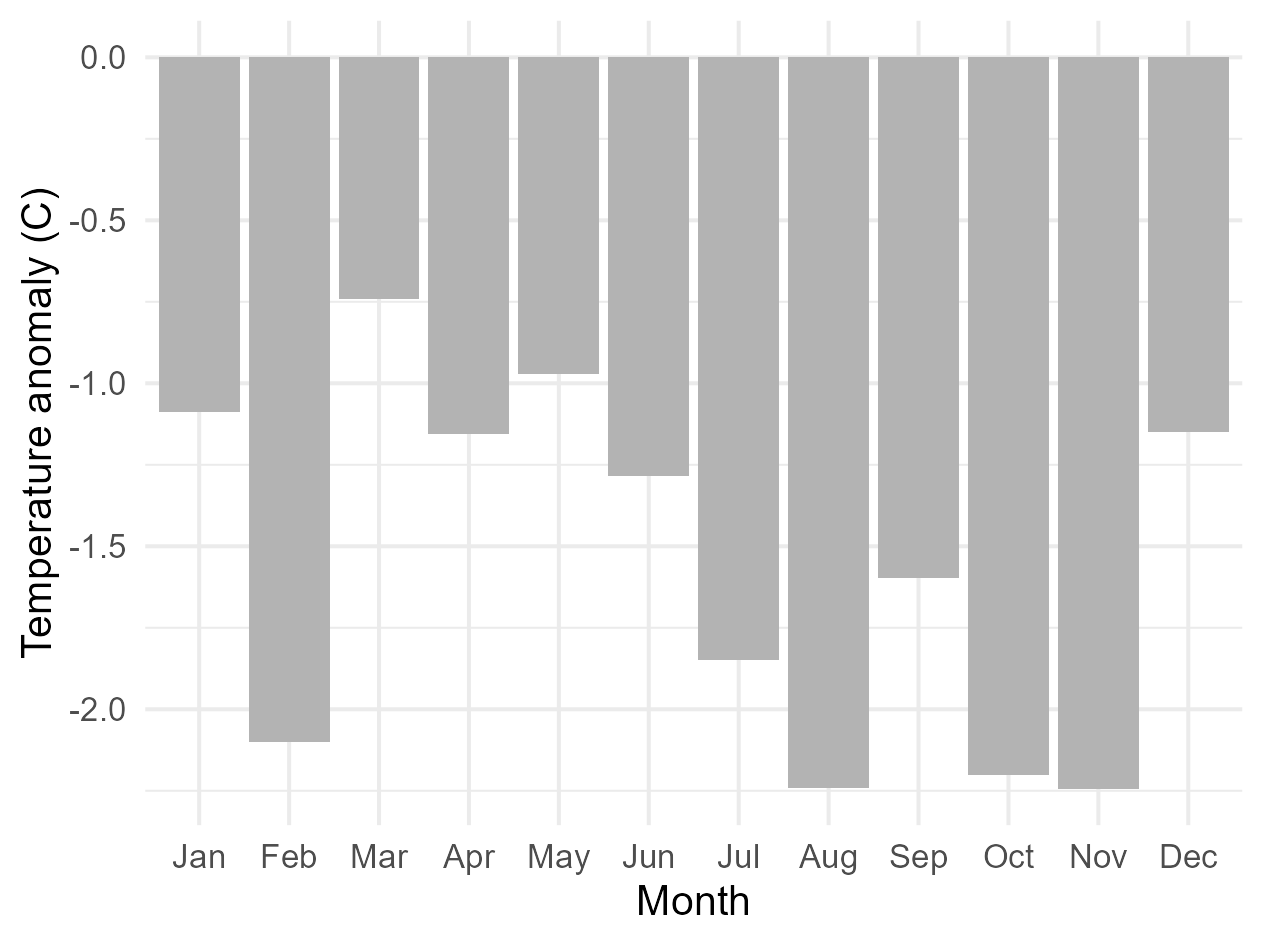

**(b)**

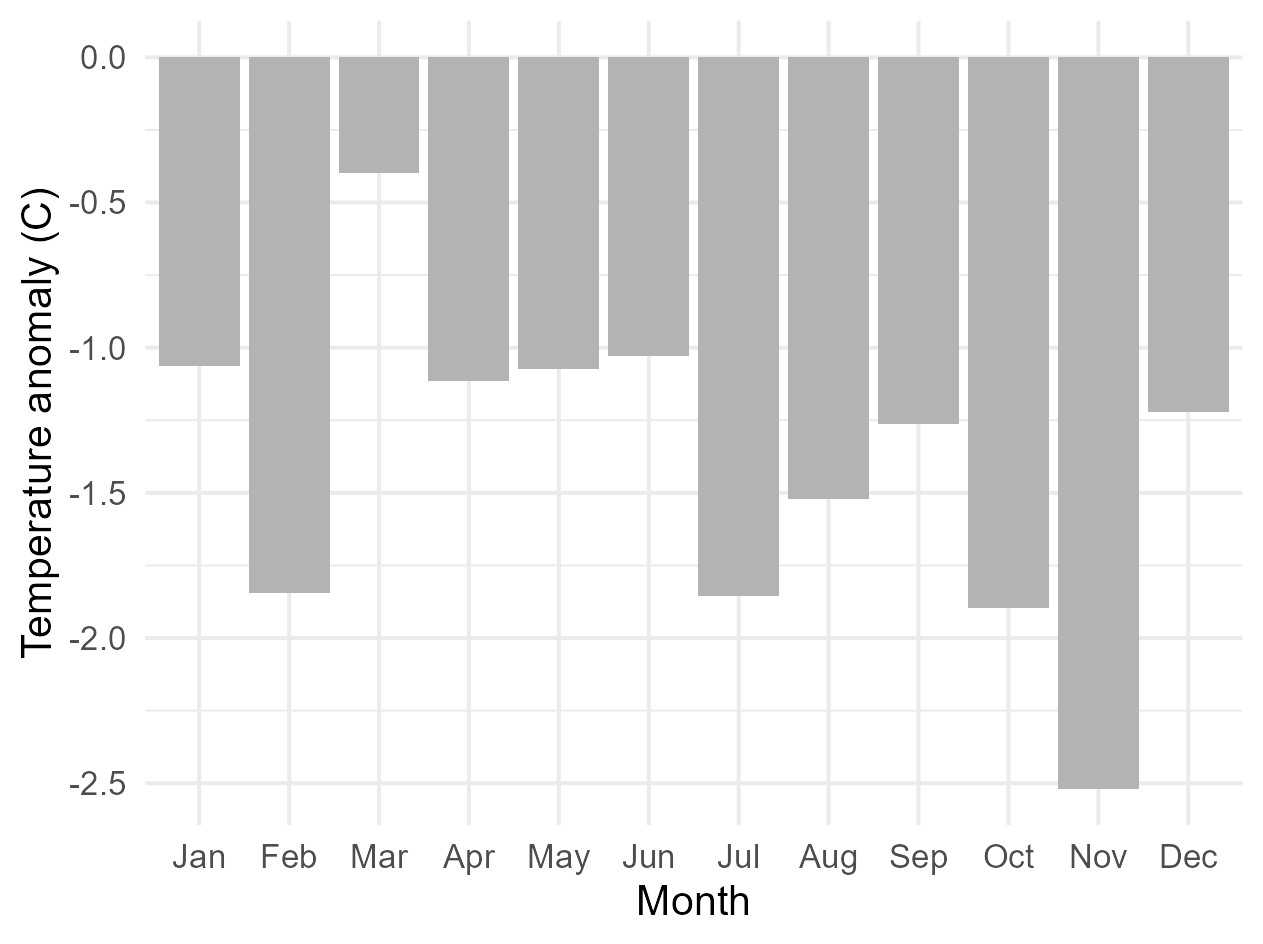

**Fig. S15.Temperature anomaly during outbreak period vs historical periods**

Bar chart of the difference in average daytime temperature by month between study period (2024-2025) vs (a) 23-year historic period (2000-2023) and (b) 5-year historic period (2018-2023). The historic period is 23 years as opposed to 30 years due to MODIS/061/MOD21CC3 data collection beginning on February 1, 2000. The temperature anomaly for each month was calculated by taking the difference between the study period mean and historical period mean. Bars are colored gray when the study period had lower average monthly temperature than the historic period and red when average monthly temperature was higher. Monthly land surface temperature from February 2000 – October 2025 was sourced from MODIS/061/MOD21C3 at a resolution of 0.05°.

| **Elevation (km) at human yellow fever case locations** | | | |
| --- | --- | --- | --- |
| **Mean** | **SD** | **Minimum** | **Maximum** |
| 1065.629 | 333.96 | 318 | 2190 |

**Table S3. Summary of elevation (km) at case locations**

Mean, standard deviation, minimum, and maximum elevation values extracted at case coordinates*.* Elevation data was sourced from WorldClim 2.1 at 30 arc seconds/1 km^2^ resolution.

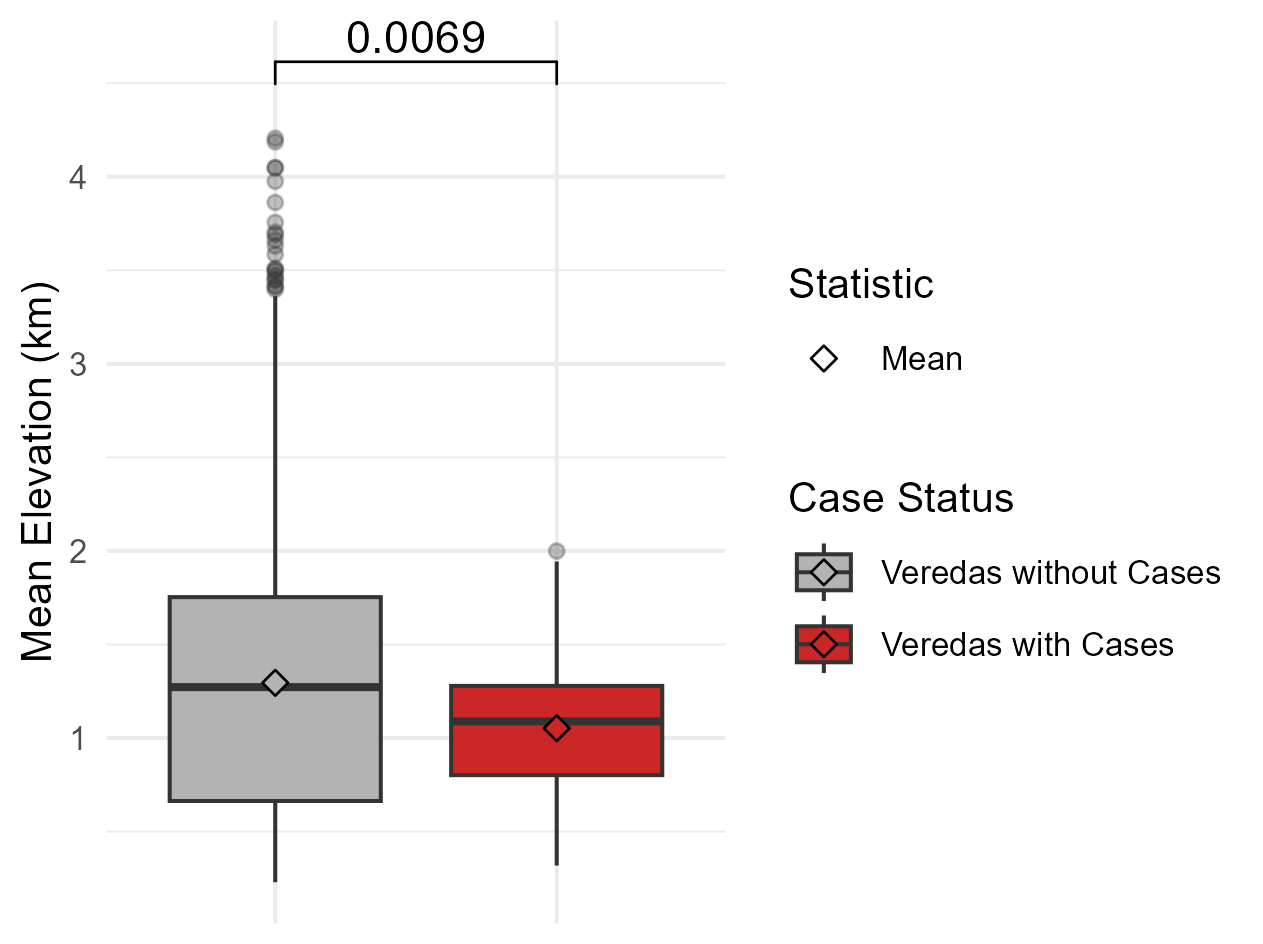

**Fig. S16. Mean elevation in veredas with cases vs those without**

Boxplot of average elevation (km) in veredas with and without human yellow fever cases in Tolima, Colombia. The central black line in each box represents group median, and the lower and upper limits of the box denote the first and third quartiles, respectively. Diamonds indicate group means. Normality was assessed using a Shapiro-Wilk test, and because elevation distributions were non-normal, a Wilcoxon rank-sum test was used to determine statistical significance between groups. The resulting p-value is displayed above the boxes. Elevation data was sourced from WorldClim 2.1 at 30 arc seconds/1 km^2^ resolution.

| **Land Cover Classification** | **Number of Cases** | **Proportion of Cases** |
| --- | --- | --- |
| Forest | 27 | 0.23 |
| Infrastructure | 4 | 0.03 |
| Mosaic of agriculture and pasture | 82 | 0.71 |
| Other non-vegetated area | 1 | 0.01 |
| Other non forest formation | 2 | 0.02 |

**Table S4. Land cover classes at human yellow fever case locations**

Number and proportion of human yellow fever cases by land cover classification in Tolima, Colombia. Land cover data from 2024 was sourced from MapBiomas Colombia at 30m^2^ resolution.

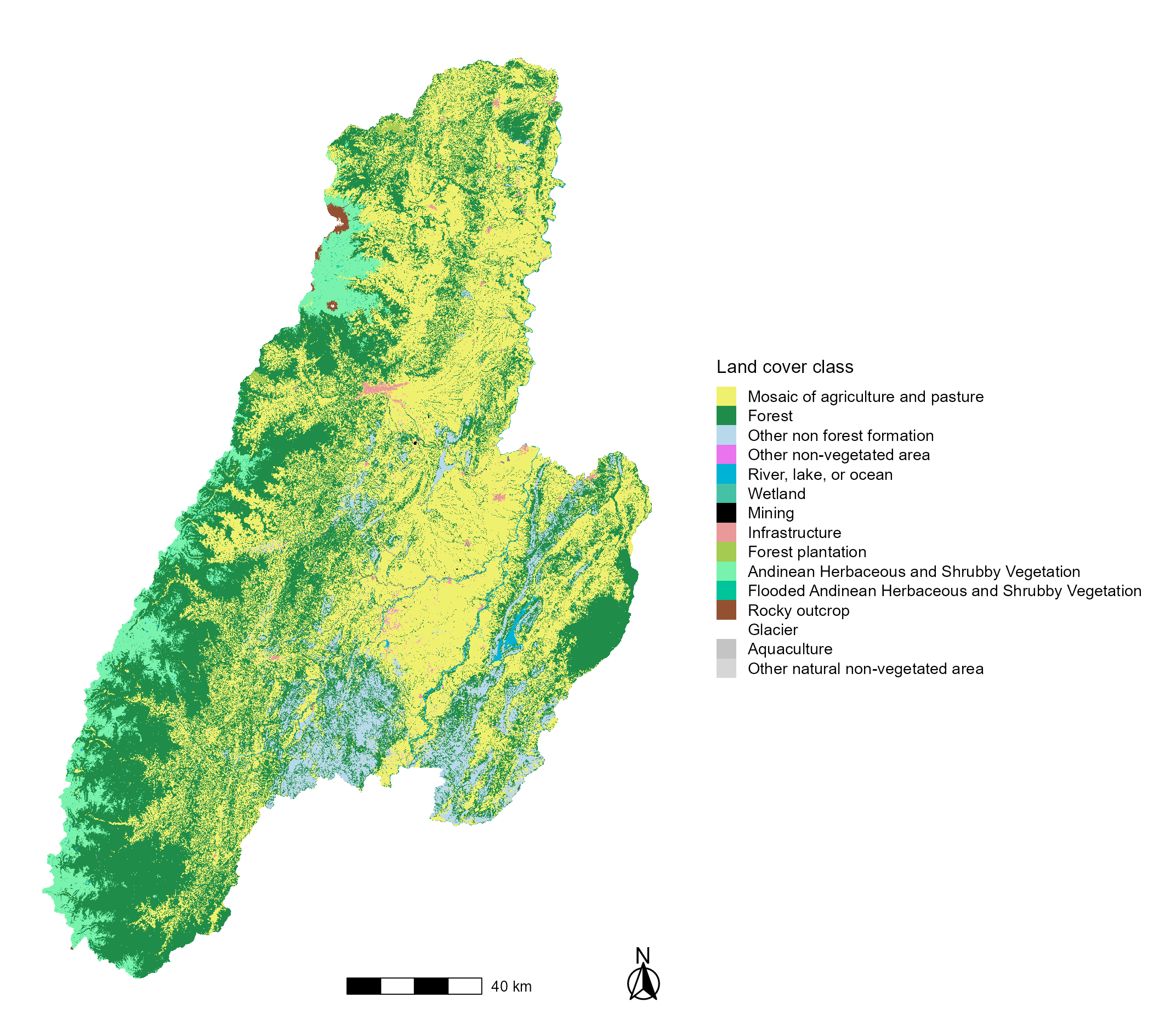

**Fig. S17. Map of landcover in Tolima, Colombia**

Map of land cover classes in Tolima, Colombia. Land cover data from 2024 was sourced from MapBiomas Colombia at 30m^2^ resolution.

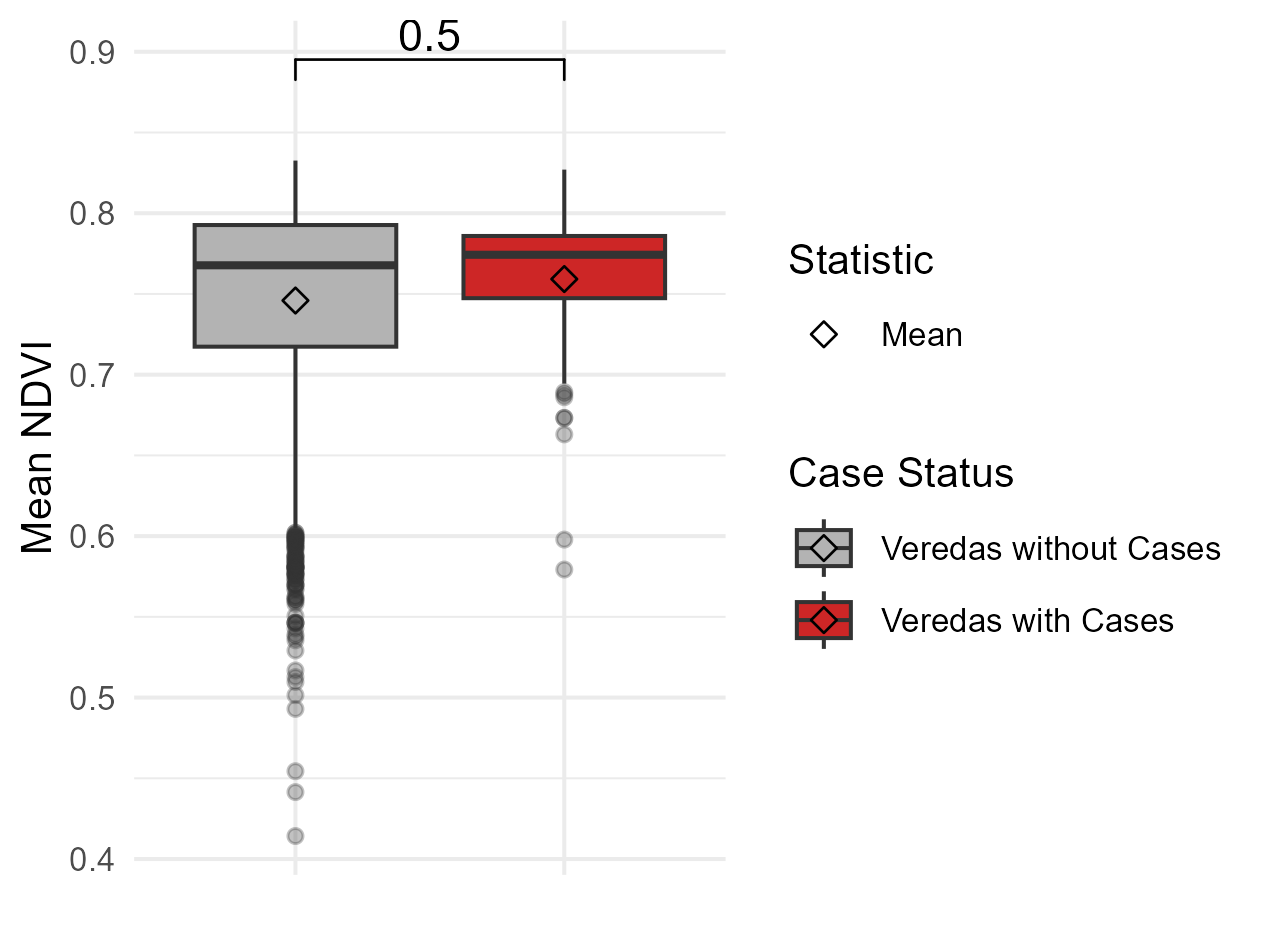

**Fig. S18. Mean Normalized Difference Vegetation Index (NDVI) in veredas with cases vs those without**

Boxplot of average NDVI values in veredas with and without human yellow fever cases in Tolima, Colombia. NDVI values range from -1 to 1, with higher values indicating denser, healthier vegetation. Mean NDVI values were extracted from raster data and averaged across all raster cells within each vereda. The central black line in each box represents group median, and the lower and upper limits of the box denote the first and third quartiles, respectively. Diamonds indicate group means. Normality was assessed using a Shapiro-Wilk test, and because NDVI distributions were non-normal, a Wilcoxon rank-sum test was used to determine statistical significance between groups. The resulting p-value is displayed above the boxes. Monthly NDVI data was sourced from MODIS/061/MOD13A3 from September 2024 – October 2025 at 1 km^2^ resolution.

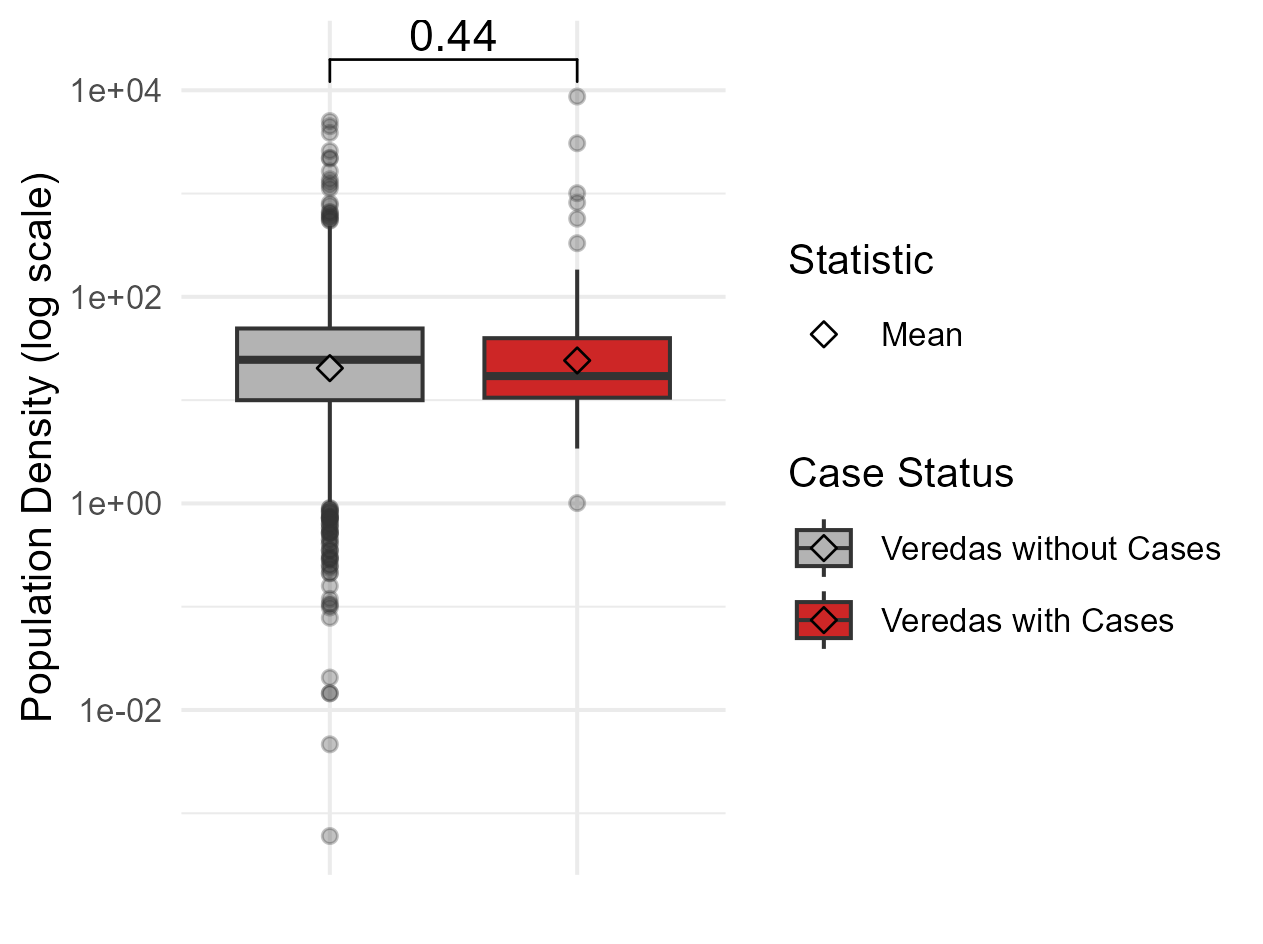

**Fig. S19. Mean population density in veredas with cases vs those without**

Boxplot of population density (people/km^2^) in veredas with and without human yellow fever cases in Tolima, Colombia and plotted on a log(10) scale. Population totals were extracted from raster data within each vereda and divided by the vereda’s total area to calculate population density. The central black line in each box represents group median, and the lower and upper limits of the box denote the first and third quartiles, respectively. Diamonds indicate group means. Normality was assessed using a Shapiro-Wilk test, and because population density distributions were non-normal, a Wilcoxon rank-sum test was used to determine statistical significance between groups. The resulting p-value is displayed above the boxes. Colombia population data in 2025 was sourced from WorldPop at 100m^2^ resolution. Population density was calculated by summing the population counts in each vereda and dividing by the vereda’s total area in km^2^.

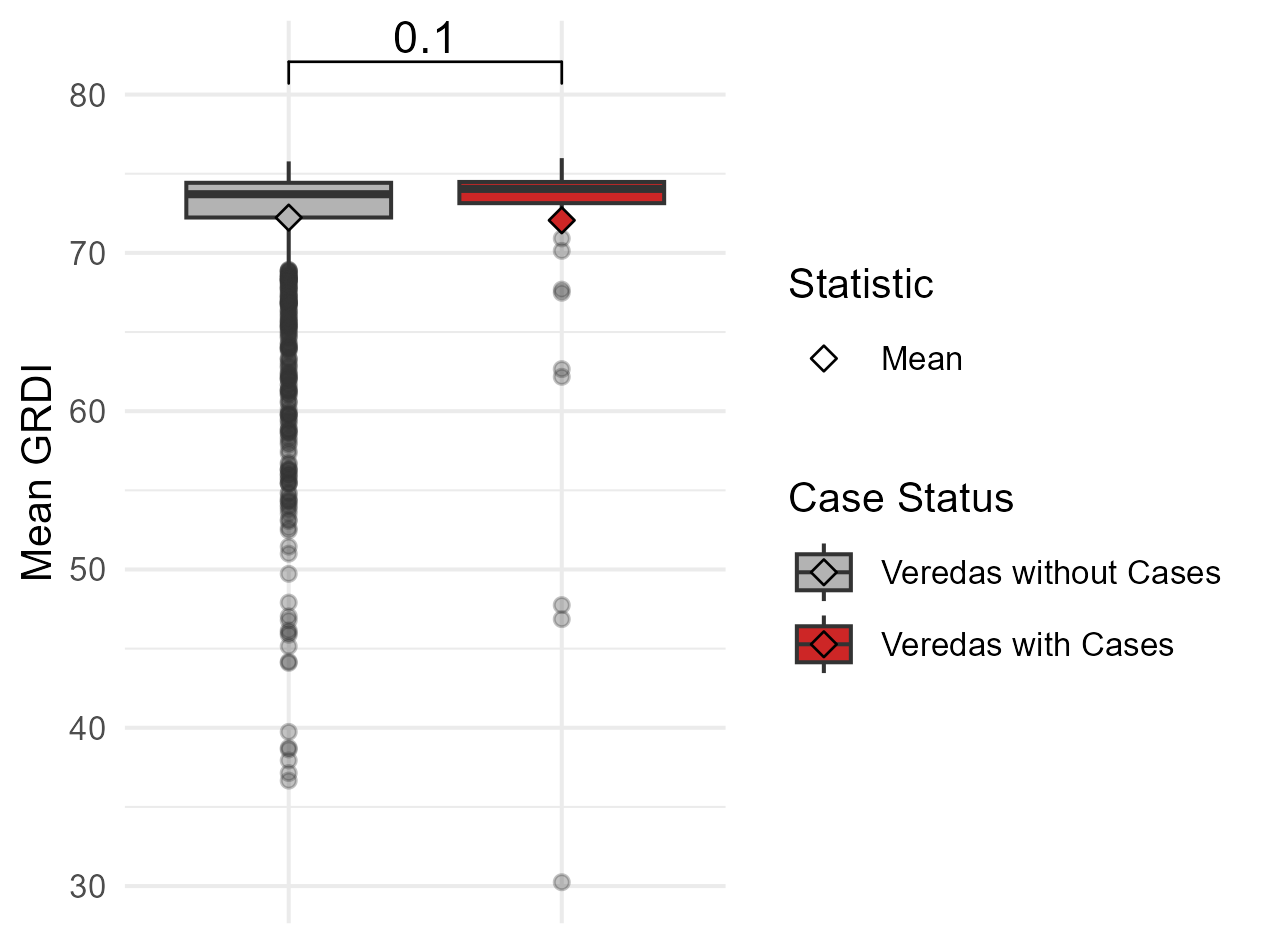

**Fig. S20. Global Gridded Relative Deprivation Index (GRDI) in veredas with cases vs those without**

Boxplot of mean Global Gridded Relative Deprivation Index (GRDI) in veredas with and without human yellow fever cases in Tolima, Colombia. The GRDI uses sociodemographic and satellite data to calculate an index of multidimensional deprivation and poverty per pixel with a range of 0-100. Data represents a temporal range from 2010-2020 and has a spatial resolution of 100 m^2^. Higher values represent greater levels of deprivation and poverty. Six dimensions are combined to define GRDI: ratio of built-up area to non-built up area, child dependency ratio (ratio between children ages 0-14 and the working population of ages 15-64), infant mortality rate, subnational human development index (assessing well-being through education, health and standard of living), intensity of nighttime lights, and the change in annual nighttime lights from 2012-2020. Twenty-one missing values were imputed using the means from spatial neighbors defined by queen contiguity. The central black line in each box represents group median, and the lower and upper limits of the box denote the first and third quartiles, respectively. Diamonds indicate group means. Normality was assessed using a Shapiro-Wilk test, and because GRDI distributions were non-normal, a Wilcoxon rank-sum test was used to determine statistical significance between groups. The resulting p-value is displayed above the boxes.

**3.8 Built-up surface**

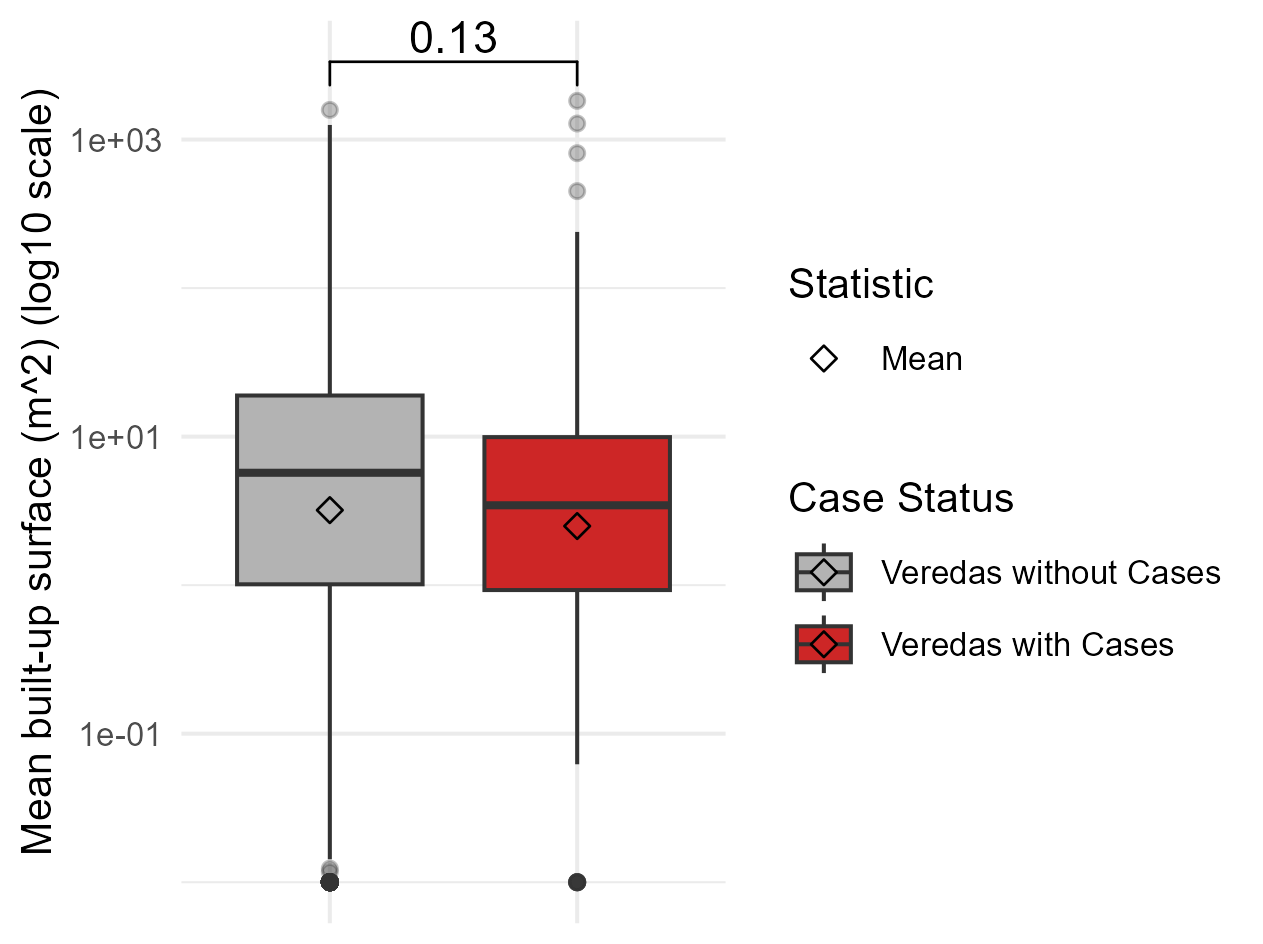

**Fig. S21. Mean built-up surface in veredas with cases vs those without**

Boxplot of average built-up surface (m^2^) values for 2025 in veredas with and without human yellow fever cases in Tolima, Colombia plotted on a log(10) scale. The central black line in each box represents group median, and the lower and upper limits of the box denote the first and third quartiles, respectively. Diamonds indicate group means. Normality was assessed using a Shapiro-Wilk test, and because built-up surface distributions were non-normal, a Wilcoxon rank-sum test was used to determine statistical significance between groups. The resulting p-value is displayed above the boxes. Built-up surface area data was sourced from Copernicus Global Human Settlement Layer at a resolution of 100 m^2^.

**6. Logistic Regression Analysis of Mortality**

|  | **Unadjusted OR** | | **Adjusted OR** | | |
| --- | --- | --- | --- | --- | --- |
| **Characteristic** | **OR** | **95% CI** | **OR** | **95% CI** | **p-value** |
| Sex |  |  |  |  |  |
| Male | 1.69 | 0.69, 4.48 | 1.80 | 0.69, 5.10 | 0.2 |
| Age (decades) | 1.43 | 1.17, 1.77 | 1.43 | 1.18, 1.78 | <0.001 |
| Abbreviations: CI = Confidence Interval, OR = Odds Ratio | | | | | |

**Table S5.**

Univariable and sex-adjusted odds ratios (ORs) and 95% confidence intervals from logistic regression models evaluating the associations between age in decades, sex, and mortality.

**7. Bayesian Hierarchical Spatiotemporal Model**

| **Model** | **IRR** | **Lower CrI** | **Upper CrI** | **DIC** | **WAIC** |
| --- | --- | --- | --- | --- | --- |
| **Model I – Fixed effects** | | | | | |
| GRDI | 2.57 | 1.92 | 3.44 | 1444.33 | 1449.38 |
| Mean temperature (3-month lag) | 1.96 | 1.43 | 2.68 | 1444.33 | 1449.38 |
| Mean precipitation (3-month lag) | 2.28 | 1.90 | 2.74 | 1444.33 | 1449.38 |
| NDVI (3-month lag) | 2.07 | 1.48 | 2.89 | 1444.33 | 1449.38 |
| **Model II – Spatial random effects** | | | | | |
| GRDI | 1.72 | 1.33 | 2.23 | 1108.74 | 1085.29 |
| Mean temperature (3-month lag) | 1.07 | 0.72 | 1.59 | 1108.74 | 1085.29 |
| Mean precipitation (3-month lag) | 1.91 | 1.59 | 2.30 | 1108.74 | 1085.29 |
| NDVI (3-month lag) | 1.41 | 1.02 | 1.96 | 1108.74 | 1085.29 |
| **Model III – Spatial Random Effects and unstructured space-time interaction** | | | | | |
| GRDI | 1.72 | 1.32 | 2.22 | 1125.91 | 1097.19 |
| Mean temperature (3-month lag) | 1.07 | 0.72 | 1.59 | 1125.91 | 1097.19 |
| Mean precipitation (3-month lag) | 1.91 | 1.59 | 2.31 | 1125.91 | 1097.19 |
| NDVI (3-month lag) | 1.41 | 1.02 | 1.96 | 1125.91 | 1097.19 |
| **Model IV – Full spatiotemporal effects** | | | | | |
| GRDI | 1.74 | 1.34 | 2.25 | 1083.64 | 1053.51 |
| Mean temperature (3-month lag) | 1.25 | 0.79 | 1.98 | 1083.64 | 1053.51 |
| Mean precipitation (3-month lag) | 2.32 | 1.62 | 3.47 | 1083.64 | 1053.51 |
| NDVI (3-month lag) | 1.37 | 0.99 | 1.89 | 1083.64 | 1053.51 |

**Table S6.**

Incidence rate ratios and 95% Bayesian credible intervals (CrI) from negative binomial models assessing the associations between yellow fever cases and GRDI (Global Gridded Relative Deprivation Index, Version 1) and lagged monthly temperature, precipitation and NDVI. Model I included fixed effects only. Model II included fixed effects and spatially structured and unstructured random effects (Besag-York-Mollie (BYM2)). Model III added an unstructured space-time interaction term (i.i.d.). Model IV was a full spatiotemporal model, additionally including a temporally structured random effect modeled as a first order random walk. Model fit was assessed using the deviance information criterion (DIC) and the Watanabe-Akaike information criterion (WAIC). The baseline model, which included only random effects and no covariates, had a DIC of 1173.31 and WAIC of 1100.33.

| **Component** | **Baseline Model**  **Variance Percentage** | **Spatiotemporal Model (Model IV)**  **Variance Percentage** |
| --- | --- | --- |
| Structured spatial (BYM2) | 90.44 | 82.97 |
| IID spatial (BYM2) | 3.94 | 6.87 |
| Temporal (RW1) | 5.36 | 4.29 |
| Space–time (IID) | 0.26 | 5.87 |

**Table S7.**

Percentage of random-effect variance attributable to structured and unstructured spatial effects, temporal effects, and space-time effects in the baseline model and the full spatiotemporal model (Model IV).
